# “MyeGPT: an AI agent for Multiple Myeloma”

**DOI:** 10.64898/2026.05.14.26353252

**Authors:** Jia Geng Chang, Alexander M. Gout, Jonathan Rodiger, Tae-Hoon Chung, George Mulligan, Wee Joo Chng

## Abstract

**Background:** Today, advancements in our understanding of cancer biology are increasingly attributed to large-scale clinical-molecular datasets. The case in point for multiple myeloma–the second-most prevalent haematological malignancy–is the CoMMpass study, a dataset with the paired clinical and sequencing data of 1,143 patients. However, the complexity of this rich dataset—with 763 clinical parameters and summary data spread across >20 files—imposes hurdles to clinician-researchers interested in making simple queries like “What percentage of patients relapse after VRD induction therapy?” or “Compare the overall survival of patients with high vs normal expression of *NSD2*”.

**Methods:** The rise of agentic AI over the past few years presents unparalleled opportunities to bridge this technical gap. We developed MyeGPT, an AI agent for clinical-molecular analysis of multiple myeloma. Based on the Reasoning-Acting (ReAct) framework, our agent converts natural language into *de novo* analyses grounded on the CoMMpass dataset, performs statistical analyses, and generates publication-quality plots. For validation, we created a benchmark of 20 calculation-intensive questions and designed two problems backed on published findings.

**Results:** MyeGPT achieves a mean reasoning-accuracy composite score of 79.4% on the internal benchmark and achieves inter-rater reliability of κ = 0.965 with human bioinformaticians. It also reproduces published findings with near perfect accuracy. We deploy the agent as a ready-to-use browser application, enabling on-the-go hypothesis validation from a smartphone.

**Conclusions:** MyeGPT demonstrates how agentic AI can eliminate the laborious scripting involved in analysing a large multi-omics dataset like CoMMpass. By increasing accessibility to a wide range of analyses from univariable statistics to transcriptome-wide hypothesis testing, MyeGPT can speed up clinical-cohort validation and hypothesis generation for multiple myeloma.

**Key points:**

1. We propose MyeGPT, a ReAct agent for the analysis and visualisation of multi-omics data of the CoMMpass study of Multiple Myeloma
2. MyeGPT obtains a reasoning-accuracy composite score of 79.4 when evaluated on a numeric response question benchmark
3. MyeGPT demonstrates high inter-rater reliability (Cohen’s κ 0.965) with human test takers on classifying functional high-risk patients
4. We used MyeGPT to reproduce analyses in the official publication of CoMMpass release IA22 related to the PR RNA-seq subtype
5. We applied MyeGPT on novel scenarios ranging from simple univariate queries, multivariate statistical testing, to transcriptome-wide multiple testing

**Biographical note:** This study is a collaboration between researchers from the laboratory of Professor Chng Wee Joo, Senior Principal Investigator, Cancer Science Institute of Singapore and the Multiple Myeloma Research Foundation, USA.

## Introduction

With 1143 newly diagnosed multiple myeloma cases, the MMRF CoMMpass Study (NCT01454297)^1,2^ stands as the third-largest cohort across any cancer type for which there is comprehensive clinical-molecular data. This publicly available resources has since enabled several landmark discoveries^2,3,4,5,6,7^. However, the large number of parameters–with 763 unique parameters from clinical tables alone–creates steep learning curve for researchers to access the CoMMpass dataset meaningfully. Various tools have been created to simplify the interaction with CoMMpass (Supplementary Table 1). Most recent is the official MMRF Virtual Lab (VLab), a graphical platform with tools such as OncoMatrix, ProteinPaint, gene expression clustering, BAM slicing, and copy number visualization. Meanwhile, MMRFBiolinks^8^ and MMRFVariant^9^ are two R libraries written for the analysis of earlier versions of CoMMpass (IA14). A limitation across these tools is their narrow choice of analyses that cannot cater to the large possibility of cross-modal analyses and hypothesis tests.

Agentic AI presents itself as a solution to the challenges of analytical tools for multi-modal datasets like CoMMpass. Developed around 2024 as a way to improve the performance of large-language models (LLMs), agentic AI extends the ability of LLMs from shallow question-answering into planning and solving workflows by incorporating the four agentic design patterns of reflection, tool-use, planning, and multi-agent collaboration^10^. General-purpose agentic AI applications for bioinformatics are on the rise and include Biomni Lab^11^, PromptBio^12^, Superbio Copilot^13^, and AI-HOPE^14^ (Supplementary Table 2).

These agents download open-source datasets, ingest user-supplied data, plan/orchestrate workflows, invoke nf-core pipelines, and support a diverse range of bioinformatics workflows such as drug discovery, pathway analysis, and single cell clustering. While these general-purpose agents are capable of genomics data analysis, the need to upload sensitive files onto their servers poses significant regulatory concerns. An additional inconvenience given the complexity of the CoMMpass dataset is the need to explicitly specify which field names to use or provide a dictionary for the field names each time the agent is initialized. Taken together, these factors create the niche for a specialized agent with pre-configured access to the CoMMpass study.

The aim of this protocol is to 1. build an AI bioinformatician to perform myeloma-related analysis and 2. to develop an evaluation dataset to ensure that MyeGPT-generated responses are factually accurate.

## Methods

### CoMMpass database

We adapted the summary files of CoMMpass into a PostgreSQL database consisting of 17 tables (overview in Table 1). Notably, it includes right-censored survival, clinical parameters, RNA-seq-derived gene expression, WGS-derived copy number, WGS- and WES-derived variant calls. Access to this PostgreSQL database is achieved through two tools: first, the ‘`Langchain SQL query`’ tool is used to run test queries and return a few rows for debugging, then the ‘`Python SQL query`’ tool is called upon to execute validated queries and write the full results to disk.

**Table 1:** Overview of the MyeGPT database. The MyeGPT database is a relational database made up of several interlinked tables derived from CoMMpass. All tables are derived from Interim Analysis 21, except for ‘`canonical_ig`’ which is obtained from IA16. Annotations: “Official”, downloaded from MMRF CoMMpass; “derived”, processed with third-party algorithms using CoMMpass data as input; Abbreviations: HGNC, HUGO Gene Nomenclature Committee.

| Table name | Description | Source |
| --- | --- | --- |
| Clinical demographics |  |  |
| per_patient | Patient-level clinical parameters | official |
| per_visit | Laboratory values taken during each visit | official |
| stand_alone_survival | Right-censored survival times | official |
| Treatment and response |  |  |
| stand_alone_trtresp | Response to various treatment lines | official |
| stand_alone_treatment_regimen | Details of various treatment lines e.g., type, duration, reason for discontinuation | official |
| Genomics |  |  |
| exome_ns_variants | Non-synonymous exome variants | official |
| canonical_ig | Recurrent IgH and Myc translocations | official |
| wgs_fish | FISH probe values and hyperdiploidy status | official |
| genome_gatk_cna | Copy number of probe segments | official |
| gatk_baf | B-allele frequencies | official |
| mutsig_sbs | Signatures of single base substitution mutations | custom |
| chromothripsis | Chromothripsis events called by ShatterSeek | custom |
| Transcriptomics |  |  |
| expr | RNA-seq counts and tpm gene expression, stored as gene-indexed arrays. | official |
| expr_metadata | A helper table for `expr` to enable lookup by sample or patient ID | official |
| Others |  |  |
| gene_annotation | Metadata and genomic coordinates of human genes | Ensembl v115 |
| hgnc_nomenclature | Aliases of HGNC-approved gene symbols and names | HGNC |
| gep_scores | Gene expression profiling scores (UAMS70, EMC92, etc.) derived from Salmon tpm estimates | custom |

The data are obtained from CoMMpass release IA22, except for canonical IgH translocation events, which was obtained from release IA16a. Most tables in the database are *official* as they closely match the official summary files. However, three tables are *derived* – chromothripsis calls (‘`chromothripsis`’), mutational signatures (‘`mutsig_sbs`’), and GEP-based risk signatures (‘`gep_scores`’). Chromothripsis was called using the Shatterseek^15^ algorithm using structural variant calls and copy number segmentation values. We adapted 4 GEP indices (UAMS70, EMC92, IFM15, MRC IX 6) to run on RNA-seq-based quantification of gene expression by mapping Affymetrix probe IDs to Ensembl gene stable IDs. SigProfiler was ran on WES data to obtain activity levels of 86 COSMIC single base substitution (SBS) signatures. We derived integer copy number statuses for the ‘`genome_gatk_cn`’ table from probe mean log ratios using the procedure detailed in another publication^16^.

### CoMMpass knowledge base

We provide MyeGPT with a 52-page documentation on how to navigate CoMMpass data (overview in Supplementary Table 3). Each page has descriptions for conceptually related field names from the same table. For example, the patient-level clinical table (‘`per_patient`’) has 7 pages: ‘`demographics`’, ‘`risk markers`’, ‘`treatment`’, ‘`treatment respons`’, ‘`death`’, ‘`miscellaneous`’, and ‘`ignor`’. These documents are bundled into a knowledge base, which is implemented as a PostgreSQL text-embedding vector store. These documents are indexed by the ‘`document search`’ tool, which retrieves the most relevant documentation pages by comparing the similarity of the query with various documentation pages in text-embedding space. As opposed to loading all pages once at initialization, we found that using Retrieval-Augmented Generation (RAG)^17^ saves tokens and avoid context decay.

### Benchmarking Procedure

#### Knowledge retrieval benchmark

As much as the CoMMpass knowledge base paginates fields by their semantic closeness, mistakes do occur during document retrieval. For example, the query term “mutation” might retrieve the mutational signatures table (‘`mutsig_sbs`’) instead of the exome mutations table (‘`exome_ns_variants`’). When the wrong documentation is retrieved, the agent searches for non-existent information in a table, wasting time and computation. Thus, text embedding quality also modulates question-answering performance. To this end, we created a test set to evaluate text embedding quality, consisting of 250 query-to-reference mappings with query terms related to multiple myeloma and CoMMpass terminology (Supplementary Table 4).

Text embedding models were tasked categorize each query into the closest existing table in the CoMMpass database e.g., the query “gene loss of function” is to the exome non-synonymous mutations table, as is the query “gene deletion” is to the gene copy number table, as is “genome-wide chromosomal rearrangements” is to the chromothripsis calls table. We compared text embedding services from four major providers: OpenAI, Google AI, Mistral, Amazon AWS. The models (‘`text-embedding-003-larg`’, ‘`gemini-embedding-001`’, ‘`mistral-embed`’ and ‘`titan-text-embed-v1`’ have embedding dimensions of 3072, 3072, 1024, and 1536 respectively. For each embedding service, we ran the document search tool to retrieve the top three documents related to the query term, ranked by PGVector vector similarity search (see ‘`document search`’ tool, Table 3). The correct document does not appear within the top three documents: the result is failure. We define N-attempt accuracy as the frequency of hits within “N” attempts.

#### Numeric response benchmark

To enable standardised evaluation of MyeGPT’s intelligence, we designed the CoMMpass Quantitative Test Set (CQTS), comprising 20 numeric response questions that requiring data-wrangling and calculation using various tables in CoMMpass. They can be categorized into three levels of difficulty. *Simple* problems involve computation of univariate statistics such as percentiles and involve only one table. *Intermediate* problems involve hypothesis testing across multiple tables. *Complex* problems designed to be arduous to solve even with the best LLMs. One example problem per difficulty level is shown in Table 2 (solutions in Supplementary File 1).

**Table 2:**
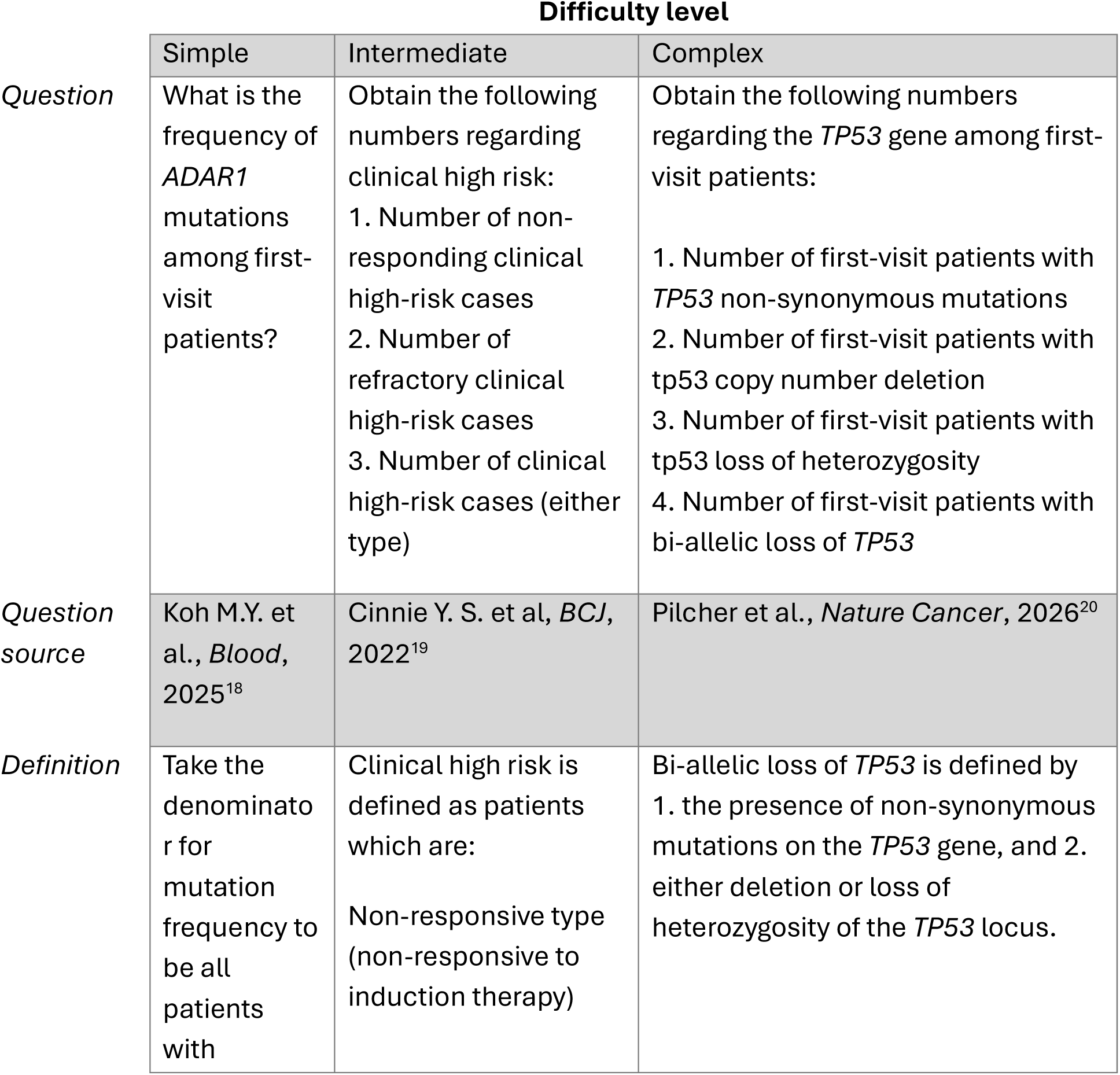
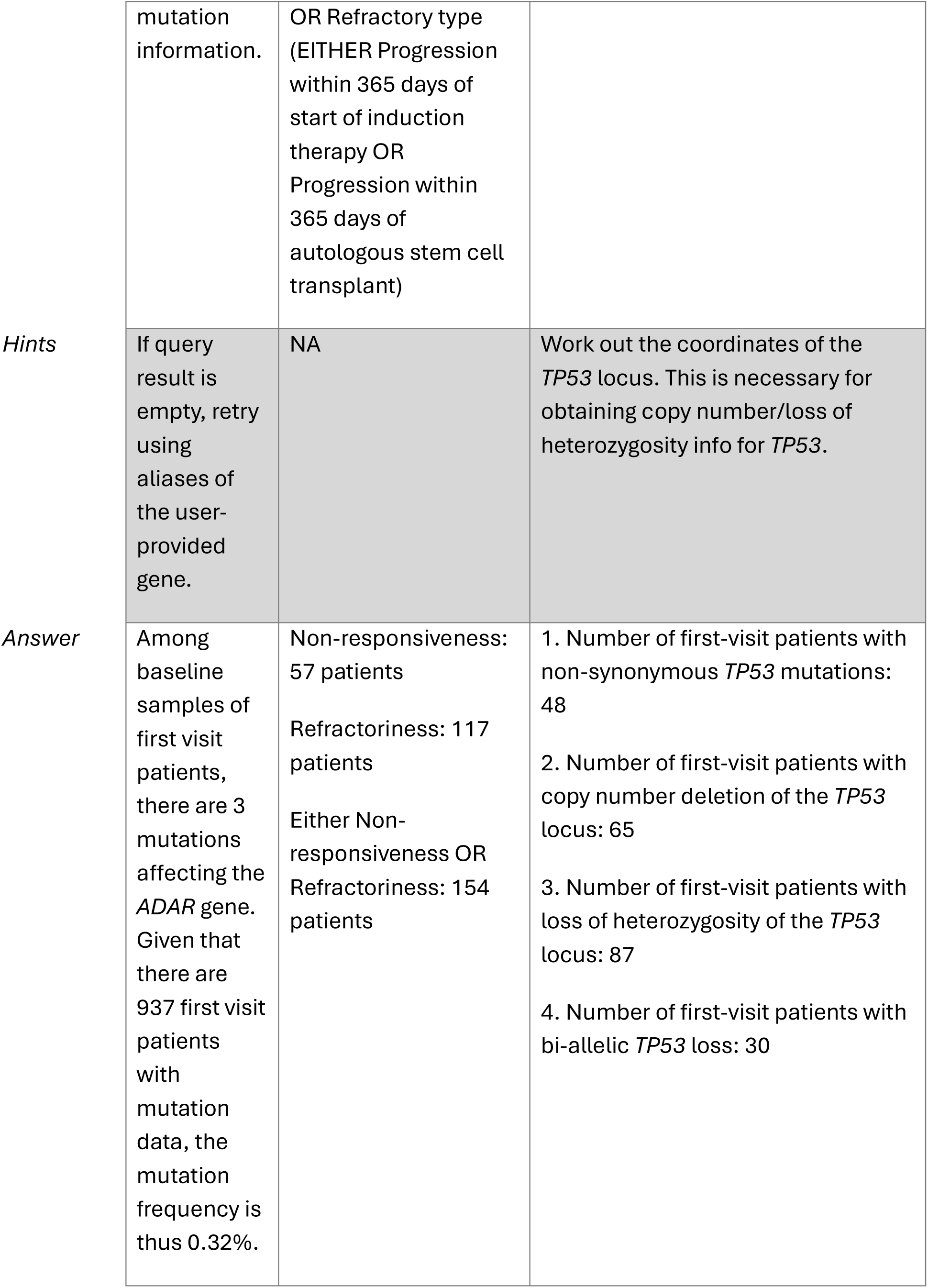
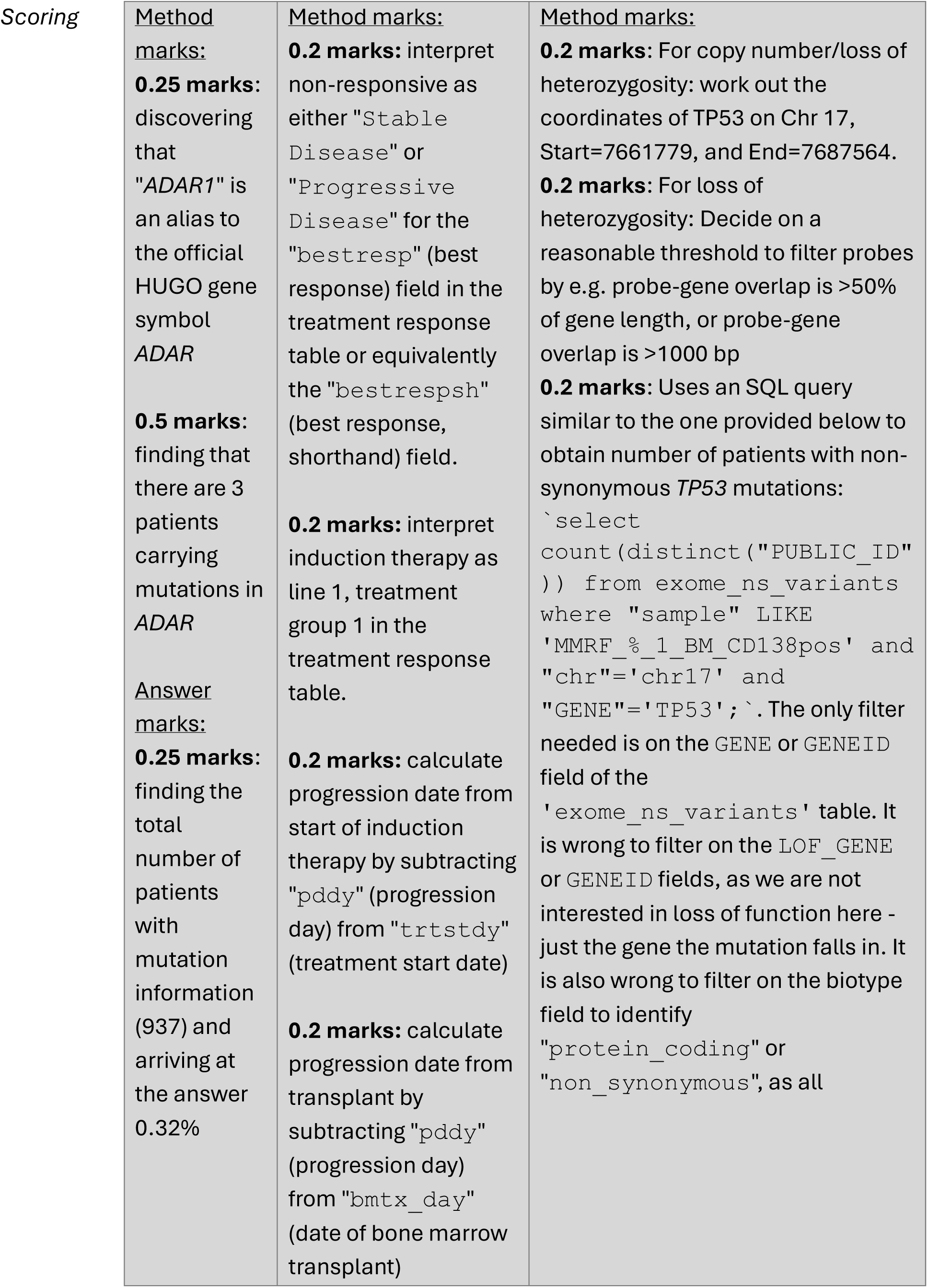
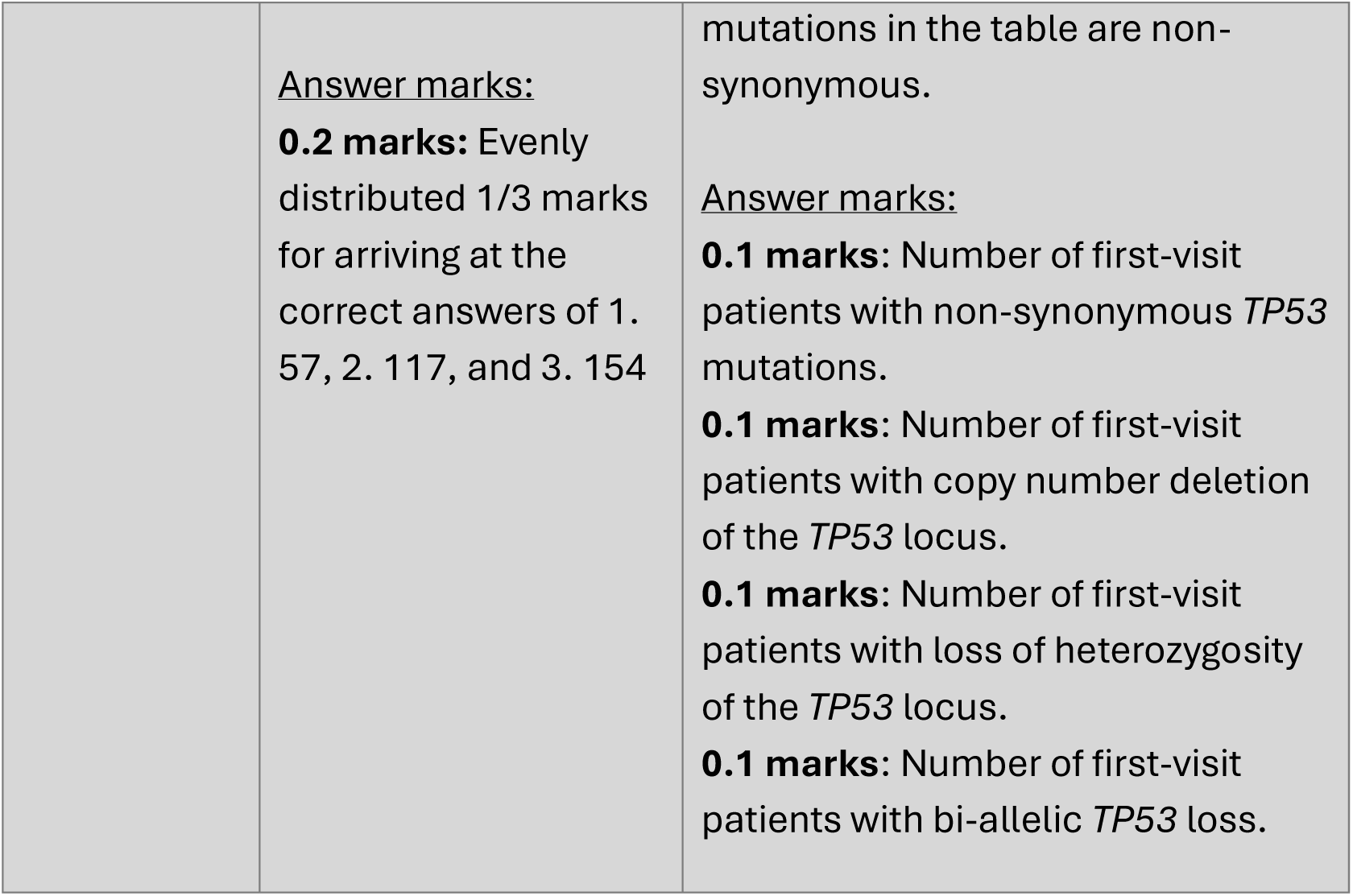
Example problems from the CoMMpass Quantitative Test Set. One question per difficulty level is selected from the 20 questions in the CoMMpass quantitative test set (CQTS). The difficulty level was determined based on the complexity of the task, the number of tables involved in the query, and the level of assistive hints or definitions if any. The score range for each question is 0-1.0. All questions are quantitative in nature e.g., find the size of a subpopulation, mutation frequency of a gene, multivariate Cox hazard ratio of a feature. The ground truth answers were obtained by the author.

|  | Difficulty level |  |  |
| --- | --- | --- | --- |
|  | Simple | Intermediate | Complex |
| *Question* | What is the frequency of *ADAR1* mutations among first-visit patients? | Obtain the following numbers regarding clinical high risk:<br>1. Number of non-responding clinical high-risk cases<br>2. Number of refractory clinical high-risk cases<br>3. Number of clinical high-risk cases (either type) | Obtain the following numbers regarding the *TP53* gene among first-visit patients:<br>1. Number of first-visit patients with *TP53* non-synonymous mutations<br>2. Number of first-visit patients with *tp53* copy number deletion<br>3. Number of first-visit patients with *tp53* loss of heterozygosity<br>4. Number of first-visit patients with bi-allelic loss of *TP53* |
| *Question source* | Koh M.Y. et al., *Blood*, 2025<sup>18</sup> | Cinnie Y. S. et al, *BCJ*, 2022<sup>19</sup> | Pilcher et al., *Nature Cancer*, 2026<sup>20</sup> |
| *Definition* | Take the denominator for mutation frequency to be all patients with | Clinical high risk is defined as patients which are:<br>Non-responsive type (non-responsive to induction therapy) | Bi-allelic loss of *TP53* is defined by<br>1. the presence of non-synonymous mutations on the *TP53* gene, and 2. either deletion or loss of heterozygosity of the *TP53* locus. |
|  | mutation information. | OR Refractory type (EITHER Progression within 365 days of start of induction therapy OR Progression within 365 days of autologous stem cell transplant) |  |
| *Hints* | If query result is empty, retry using aliases of the user-provided gene. | NA | Work out the coordinates of the *TP53* locus. This is necessary for obtaining copy number/loss of heterozygosity info for *TP53*. |
| *Answer* | Among baseline samples of first visit patients, there are 3 mutations affecting the *ADAR* gene. Given that there are 937 first visit patients with mutation data, the mutation frequency is thus 0.32%. | Non-responsiveness: 57 patients<br>Refractoriness: 117 patients<br>Either Non-responsiveness OR Refractoriness: 154 patients | 1. Number of first-visit patients with non-synonymous *TP53* mutations: 48<br>2. Number of first-visit patients with copy number deletion of the *TP53* locus: 65<br>3. Number of first-visit patients with loss of heterozygosity of the *TP53* locus: 87<br>4. Number of first-visit patients with bi-allelic *TP53* loss: 30 |

Scoring
|  |  |  |
| --- | --- | --- |
| <u>Method marks:</u><br>**0.25 marks:** discovering that "ADAR1" is an alias to the official HUGO gene symbol ADAR<br>**0.5 marks:** finding that there are 3 patients carrying mutations in ADAR<br><u>Answer marks:</u><br>**0.25 marks:** finding the total number of patients with mutation information (937) and arriving at the answer 0.32% | <u>Method marks:</u><br>**0.2 marks:** interpret non-responsive as either "Stable Disease" or "Progressive Disease" for the "bestresp" (best response) field in the treatment response table or equivalently the "bestrespsh" (best response, shorthand) field.<br>**0.2 marks:** interpret induction therapy as line 1, treatment group 1 in the treatment response table.<br>**0.2 marks:** calculate progression date from start of induction therapy by subtracting "pddy" (progression day) from "trtstdy" (treatment start date)<br>**0.2 marks:** calculate progression date from transplant by subtracting "pddy" (progression day) from "bmtx_day" (date of bone marrow transplant) | <u>Method marks:</u><br>**0.2 marks:** For copy number/loss of heterozygosity: work out the coordinates of TP53 on Chr 17, Start=7661779, and End=7687564.<br>**0.2 marks:** For loss of heterozygosity: Decide on a reasonable threshold to filter probes by e.g. probe-gene overlap is >50% of gene length, or probe-gene overlap is >1000 bp<br>**0.2 marks:** Uses an SQL query similar to the one provided below to obtain number of patients with non-synonymous TP53 mutations:<br>``select count(distinct("PUBLIC_ID" )) from exome_ns_variants where "sample" LIKE 'MMRF_%_1_BM_CD138pos' and "chr"='chr17' and "GENE"='TP53';`. The only filter needed is on the GENE or GENEID field of the 'exome_ns_variants' table. It is wrong to filter on the LOF_GENE or GENEID fields, as we are not interested in loss of function here - just the gene the mutation falls in. It is also wrong to filter on the biotype field to identify "protein_coding" or "non_synonymous", as all` |
|  | <u>Answer marks:</u><br>**0.2 marks:** Evenly distributed 1/3 marks for arriving at the correct answers of 1. 57, 2. 117, and 3. 154 | mutations in the table are non-synonymous.<br><u>Answer marks:</u><br>**0.1 marks:** Number of first-visit patients with non-synonymous *TP53* mutations.<br>**0.1 marks:** Number of first-visit patients with copy number deletion of the *TP53* locus.<br>**0.1 marks:** Number of first-visit patients with loss of heterozygosity of the *TP53* locus.<br>**0.1 marks:** Number of first-visit patients with bi-allelic *TP53* loss. |

Each problem is divided into three sections – the query, definitions (optional), and hints (optional). A set of scoring rules is provided for every question. Definitions are provided for subjective concepts, like “clinical high risk”. “responder”, or “high-risk translocations”. Hints are provided where necessary to guide the agent towards a desired approach when multiple valid approaches exist. LLM responses were automatically scored using LLM-as-a-Judge (Supplementary Text 1) using OpenAI’s GPT-5 mini, then manually vetted for any mistakes in the scoring.

Besides correctness, we allocated marks for reasoning because agent may A. provide incorrect answer, B. exceed the 50-step recursion limit or C. exceed the 600s time limit. The user can prompt the agent to continue thinking for scenarios B and C, but this is not possible in single-turn evaluation setting. Thus, method marks serve to distinguish the performance among test takers whose final answer is incorrect. We refer to the resulting score the *Reasoning-Accuracy Composite* (RAC) to reflect how both methodology and correctness are considered in the mark allocation.

For ranking LLM performance on single-turn benchmark, our primary metric is the mean RAC score across all questions in CQTS (mRAC; range 0.00-1.00). A secondary metric we use is the Net Promoter Score (NPS), calculated by the number of full marks responses subtracted by number of zero marks responses. A high or positive NPS indicates a tendency to provide perfect answers; a low or negative NPS indicates a tendency to completely fail to answer questions.

### Reproducing Chung’s FHR analysis

We initially tried one-shot prompting MyeGPT with the task of classifying functional high risk (FHR) cases. This proved infeasible due to the task complexity, as no model obtained a satisfactory answer. We resorted to a “divide and conquer” approach which proved more effective. We split the problem into the identification of TP53 double-hit patients, t(4;14)/1q21 gain patients, identification of clinical high-risk patients, and identification of FHR cases. Using a multi-turn prompting style also gave the opportunity for MyeGPT to clarify its understanding, or for the user to course-correct MyeGPT’s workflow.

The choice of LLMs evaluated here was the top five LLMs from the Numeric response benchmark. Two authors acted as human test takers for the multi-turn benchmark: J.G.C., who attempted using Python (Supplementary File 2), and T.H.C, who used R. Rather than using human responses as the ground truth, we were interested in the inter-agreement between the LLM- and human responses. This better reflects the inherently ambiguous nature in risk classification of the newly diagnosed multiple myeloma; disagreement also exists between humans. We chose Cohen’s κ or inter-rater reliability is given by:

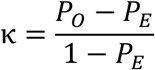

where *P*_*O*_ is the proportion of observed agreement (actual agreements divided by total items) and *P*_*E*_ is the proportion of agreement expected by chance, accounting for random agreement. κ = 1 indicates complete agreement between two raters, κ = 0 indicates agreement on pure chance, and κ < 0 indicates a real tendency for disagreement.

### Reproducing Skerget’s PR subtype analysis

Patient RNA subtype and biobank information were obtained from Supplementary Table 1 of Skerget et al. 2024^2^. We used the pyComBat^21^ package for batch correction on 714 baseline bone marrow samples by bio-bank. We supplied batch-corrected tpm values to MyeGPT to recalculate the formula for Bergsagel Proliferation index as the arithmetic mean of log2(TPM+)-transformed expression of the following 12 genes: *TYMS, TK1, CCNB1, MKI67, KIAA101, KIAA0186, CKS1B, TOP2A, UBE2C, ZWINT, TRIP13* and *KIF11*. MyeGPT recognised that *KIAA101* and *KIAA0186* were aliases to HGNC-approved symbols *PCLAF* and *GINS1*. It also detected a misnaming of *KIAA0101* as *KIAA101* in the source publication. We designed prompts (Supplementary Text 2) to reproduce the three analyses and Figure 5 in Reproducing Skerget’s PR subtype analysis.

### Applying MyeGPT to novel scenarios

We used a cost-effective version of MyeGPT instantiated with OpenAI’s `GPT-5 mini` as the LLM and `text-embedding-003-large` for knowledge retrieval. Each analysis was performed on a fresh initialisation of the agent with no past conversations (see prompts in Supplementary Text 3). MyeGPT’s responses to the analyses were manually vetted by J.G.C. to be accurate. While the seven of the eight analyses are based on internal CoMMpass data, the complex-difficulty analysis of Figure 7g relies on user-provided patient lists as part of identifying mutations enriched in a subpopulation. This serves to test the agent’s ability to ingest new information.

For the clinical Cox PH model (Figure 7e), MyeGPT was asked to standardize the following normally distributed variables: serum calcium, creatinine, LDH, β2M levels, and age, while leaving ECOG as an ordinally encoded variable (stage 0 to 4).

For analysis of mutated genes in FHR and SR patients (Figure 7g), we instructed MyeGPT to exclude immunoglobulin or chromosome Y genes, and genes with mutation recurrence of ≤3 in the FHR plus SR groups combined. Independent Fisher’s exact tests were run on the 16 resulting genes on all N=1143 patients. Patient lists for FHR and SR were provided by J.G.C. as part of the solution for multi-turn benchmark.

Kruskal-Wallis test was performed for difference in *NEDD4-1* expression (Figure 7c) between good-response (N=77) vs poor-response (N=47) and partial-response (N=171) vs poor-response, and for the analysis of differentially expressed genes by bortezomib response (Figure 7h) on 19,848 protein coding genes of N=504 responders and N=24 non-responders and corrected for multiple testing using the Benjamini-Hochberg procedure.

## Results

### MyeGPT: an AI agent for multiple myeloma

We develop MyeGPT (Myeloma Generative Pretrained Transformer; Figure 1), an AI bioinformatician for multiple myeloma. MyeGPT converts natural language queries about multiple myeloma into *de novo* analyses on the CoMMpass dataset. Central to the agent is the LLM prompt (Supplementary Text 4). This prompt frames the agent as a myeloma research assistant whose actions are to consult documentation, query its database, analyse SQL results, and visualize results. We developed MyeGPT because of the shortcomings of general-purpose agents with respect to analysing CoMMpass data; when faced with a question specific to CoMMpass study, general-purpose agents such as ChatGPT, Anthropic Claude, and Perplexity will attempt to access CoMMpass data from public portals like cBioPortal and GDC Data Portal, fail at doing so because CoMMpass is controlled-access, and urge the user to upload their own copy of CoMMpass. On the other hand, MyeGPT comes pre-loaded with CoMMpass data and is equipped with tools (Table 3) to interact with the dataset.

**Table 3.** The main tools that MyeGPT has access to. Auxiliary tools include a file path generator tool for saving/retrieval of results and graphs to disk, a HTML graph embedder tool, a gene metadata lookup tool, and a gene name conversion tool.

| **Tool name** | **Description** | **Input** | **Output** |
| --- | --- | --- | --- |
| *Document search* | Search for the most relevant reference manual of a table within the database based on the specified term.<br>The RAG-as-a-tool configuration allows it to be repeatedly called to serve complex user queries involving pulling data from different tables.<br>Depends on Pgvector<sup>1</sup> | 1. A query string related to myeloma<br>2. An integer K<br>Examples:<br>1. “missense mutation”<br>2. “overall survival”<br>3. “treatment with dexamethasone” | Reference manuals of the top K articles that most closely relates to the query<br>Examples:<br>1. {docs for the exomic variants table}<br>2. {docs for survival data table}<br>3. {docs for the treatment regimen table} |
| *Langchain SQL query* | Constructs syntactically correct SQL queries from natural language instructions, evaluates the queries, and retrieves results with an additional `LIMIT 3` clause.<br>Depends on QuerySQLDatabaseTool from Langchain-Community | An AI-generated SQL query that is unvalidated. | If query is successful, a result limited to 3 rows.<br>If the query fails an error message is returned. |
| *Python SQL query* | Opens a python connection to the database and executes SQL queries without the `LIMIT 3` clause and saves the results to disk<br>Depends on Psycopg<sup>2</sup> | A valid SQL query that has been tested before with the `Langchain SQL query` tool<br>A csv file path to save the results to. | A .csv file containing the SQL query results. |

<sup>1</sup> Pgvector: an open-source Postgres extension for similarity search
<sup>2</sup> Psycopg: an open-source PostgreSQL driver for Python

|  |  |  |  |
| --- | --- | --- | --- |
| *Gene-level copy number* | Runs the maximum overlapping segment algorithm to infer integer copy number for a genomic region<br>Depends on `Psycopg` | A genomic coordinate in the form (chromosome, start, end) | The integer copy-number level (-2, -1, 0, +1, +2) for all patients with copy number data, for the coordinates |
| *Cox model base data* | Retrieve a template dataset for Cox PH regression analysis.<br>Depends on `Psycopg` | The endpoint; either OS or PFS | A table with right-censored survival, age, sex, ISS stage of all patients in CoMMpass |
| *Python REPL3* | An interactive python shell for plotting, statistical analysis, and calculations.<br>Depends on `PythonREPLTool` from Langchain-Experimental | An AI-generated python script | Dynamic; can be a plot, a text file, or standard output |

**Figure 1.**
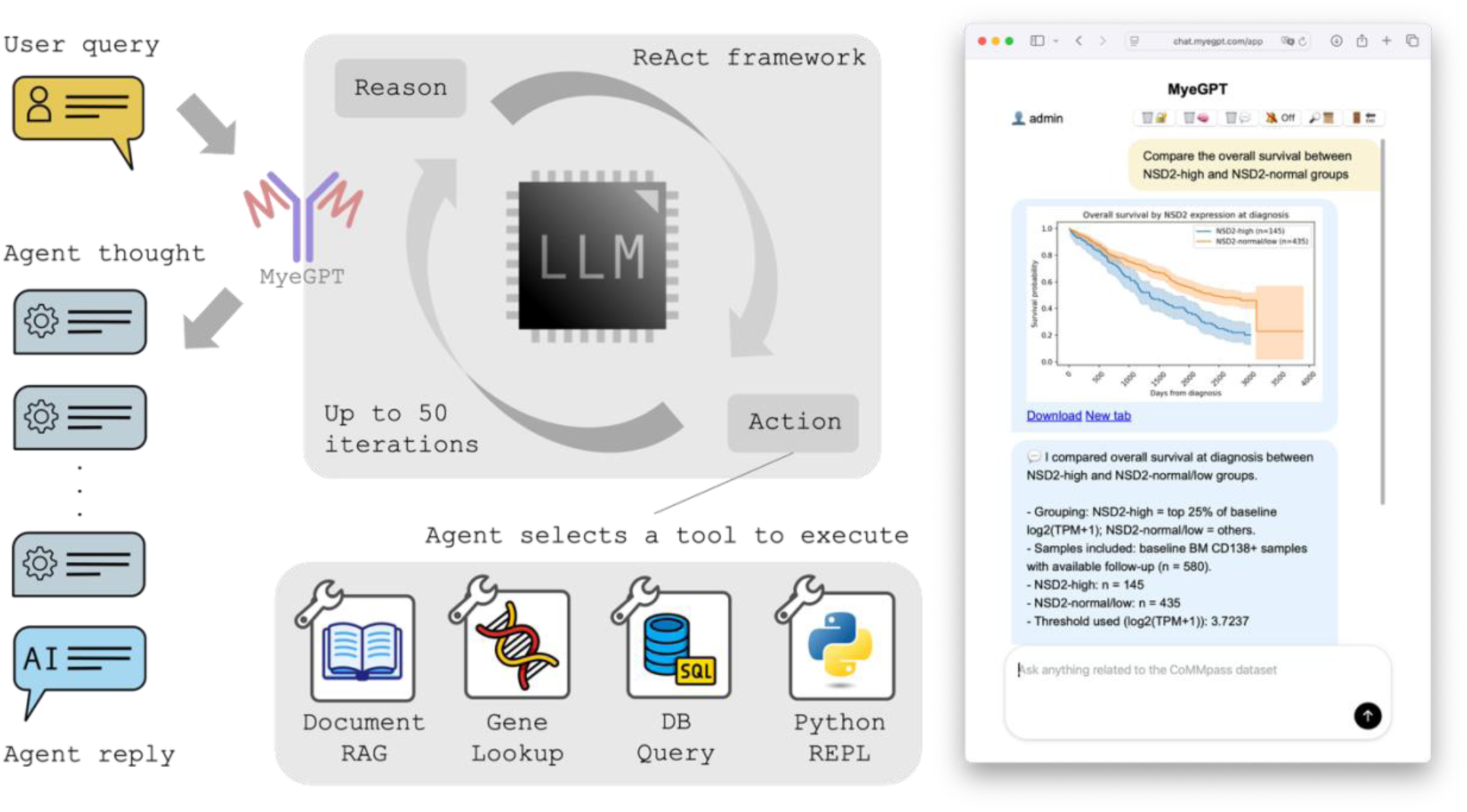
Overview of MyeGPT. Left, a simplified schematic of MyeGPT. The user query is passed into the ReAct agent, which is backed by a LLM with access to tools including the CoMMpass knowledge base among others. Right, a screenshot of the smartphone-compatible web application, showing its conversational and visualization functions.

MyeGPT employs the ReAct paradigm^22^, in which an agent undergoes alternating steps of reason and action in response to a question such as “Compare the overall survival between *NSD2*-high and *NSD2*-normal groups” (Figure 1). In the “Reason” step, the reasoning ability the LLM is called upon to create, maintain or modify the plan of action e.g. I. Find where the data is stored, II. Define cut-offs for “high” vs “normal” *NSD2* expression, III. Gather the data, IV. plot Kaplan-Meier curves. In the “Action” step, the LLM in MyeGPT calls a “tool” decided by the Reason step. With tools, the agent can interact with the external environment outside of its containerized filesystem, such as 1. Generating and running an SQL query on the CoMMpass documentation, 2. Generating and running a query the CoMMpass database, and 3. Generating and running a Python script to plot KM curves. On the next “Reason” step, the agent analyses the results of the tool and updates its plan using the knowledge gained from its interaction with the external environment via tools. Upon completion of the plan or after 50 iterations, it exits the loop and returns an AI response.

MyeGPT uses RAG-as-a-tool to enhance accuracy of its responses. The CoMMpass knowledge base (overview in Supplementary Table 3) connects user queries to specific tables of the CoMMpass database. For example, in response to the query “Calculate the median OS with high APOBEC activity”, the agent first recognizes it needs to search documentation for the term “APOBEC”. Next, the agent retrieves documentation for the mutational signatures table, where it is mentioned that signatures SBS2 and SBS13 are APOBEC-induced. In addition, this process is repeated for OS, pulling out the documentation for the survival data table. Together, these mechanisms improve the reliability and accuracy of its answers.

### Systematic benchmarking of MyeGPT

Being powered by generative AI, the reliability of MyeGPT’s responses is of major concern. We tailored a two-step approach to assess its intelligence. Step One is to optimise MyeGPT on a suite of internal benchmarks (§Knowledge retrieval benchmark, §Numeric response benchmark). Step Two is to deploy the optimised version of MyeGPT to recapitulate known findings (§Reproducing Chung’s FHR analysis, §Reproducing Skerget’s PR subtype analysis). Having confirmed its reliability, we then explore the range of analyses which MyeGPT can assist researchers in (§Applying MyeGPT to novel scenarios).

#### Knowledge retrieval benchmark

First, we measured document classification accuracy as it is the primary determinant of embedding quality. Google’s `gemini-embedding-001` and OpenAI’s `text-embedding-003-large` models achieved comparable first-attempt accuracy (68% and 67% respectively, Figure 2a). Mistral’s 1024-dimensional `mistral-embed` model and Amazon’s 1536-dimensional `titan-embed-text-v1` model performed poorer at 59% and 53% respectively. We also consider two-attempt accuracy, given that >30% of first attempts are incorrect. Again, the embedding function with best two-attempt accuracy was Google’s `gemini-embedding-001` at 0.84 followed by OpenAI’s `text-embedding-003-large` at 0.82. When we measured the Silhouette scores of the embeddings, where it was confirmed that Google’s `gemini-embedding-001` had highest scores on both Euclidean and cosine distance metrics (0.127 and 0.075; Figure 2b).

Next, we visualized the clustering of query terms by the document they are expected to match to serve as another indicator of embedding quality. The N-dimensional embedding vectors were projected to two dimensions using t-SNE then labelled by their reference table. The 3072-dimensional embeddings were more cohesively grouped by reference document label (Figure 2c-d) in comparison to the 1024-dimensional Mistral `mistral-embed` and 1536-dimensional Amazon `titan-text-embed-v1` (Figure 2e-f). Together, our analyses indicated that 3072-dimensional embeddings were superior—with little difference between the top-performers `gemini-embedding-001` and `text-embedding-003-large`. As we encountered technical issues with the API service of Google’s `gemini-embedding-001`, we turned to the equally performant OpenAI’s `text-embedding-003-large` as our final choice.

**Figure 2:**
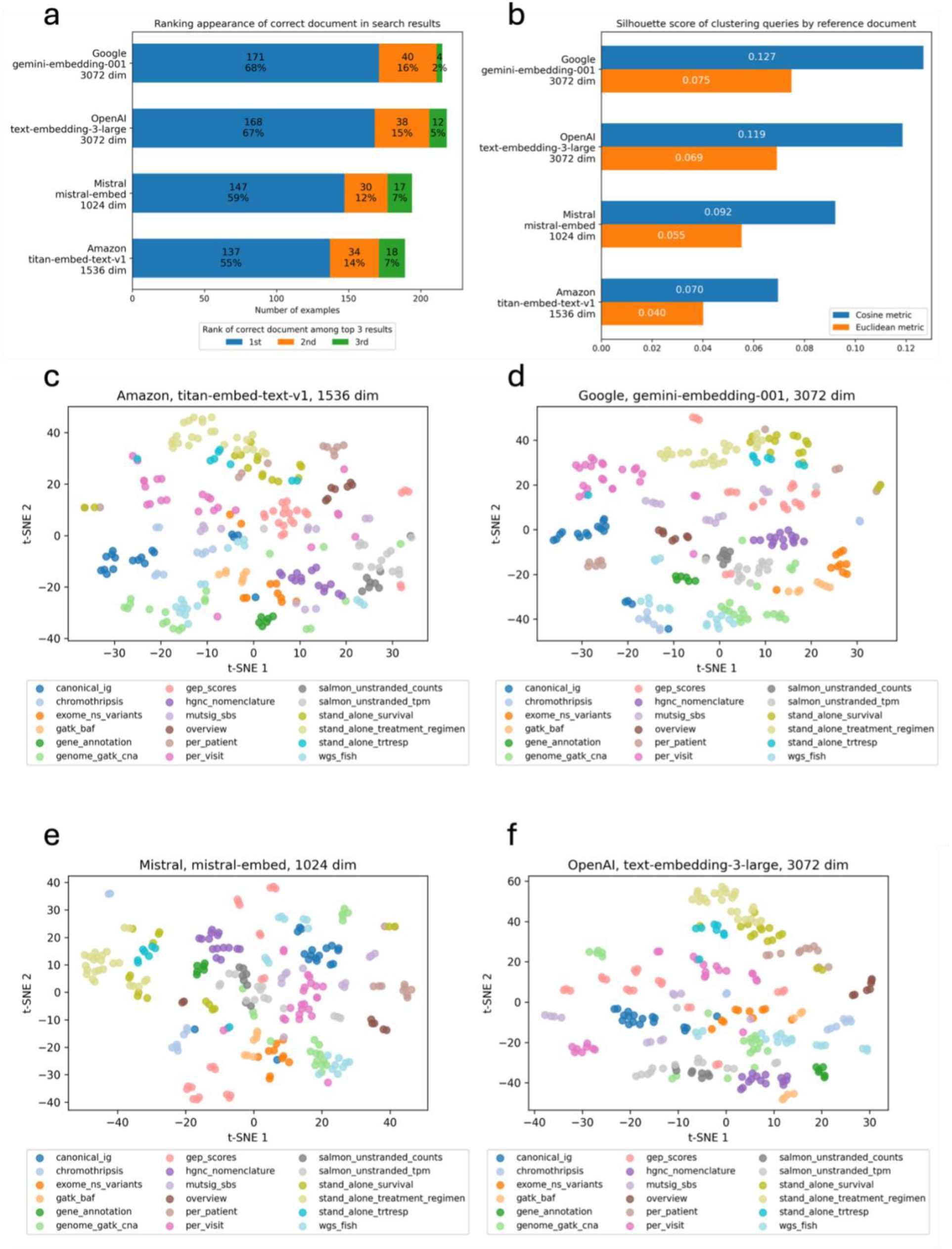
Performance of text-embedding services on 250 myeloma-related terms. (a) Attempts required to retrieve the correct table given a query, binned into first-, second-, third-, and >3 attempts. (b) Silhouette scores (cosine or Euclidean metric) by embedding service when queries are clustered based on the correct documentation. (c), (d), (e), (f) Two-dimensional tSNE projections of query embeddings, one for each neural embedding algorithm. Query embeddings are labelled by the most relevant topic for that term.

#### Numeric response benchmark

We compared different LLMs in their answer quality of CQTS questions. mRAC and NPS were used as primary and secondary metrics. We observed perfect rank-order agreement between mRAC and NPS was observed (Figure 3a). Among the LLMs tested, a clear trend of superiority of models from Anthropic and OpenAI emerged, with the top seven models hailing from these model families (Figure 3a). The flagship Anthropic Claude Opus 4.5 was the best performing among all LLMs with a mRAC of 0.794 and NPS of +11 (Table 4 row 1). Anthropic’s Claude Opus 4.5 was also the only LLM which avoided any zero-score responses. Surprisingly, the top performing model among OpenAI models was the budget friendly GPT-5 mini, with mRAC and NPS of 0.681 and +6 (Table 4 row 3) in comparison to the parametrically larger GPT-5.1 and GPT-5.2 models (Table 4, rows 4 & 6).

**Table 4:**
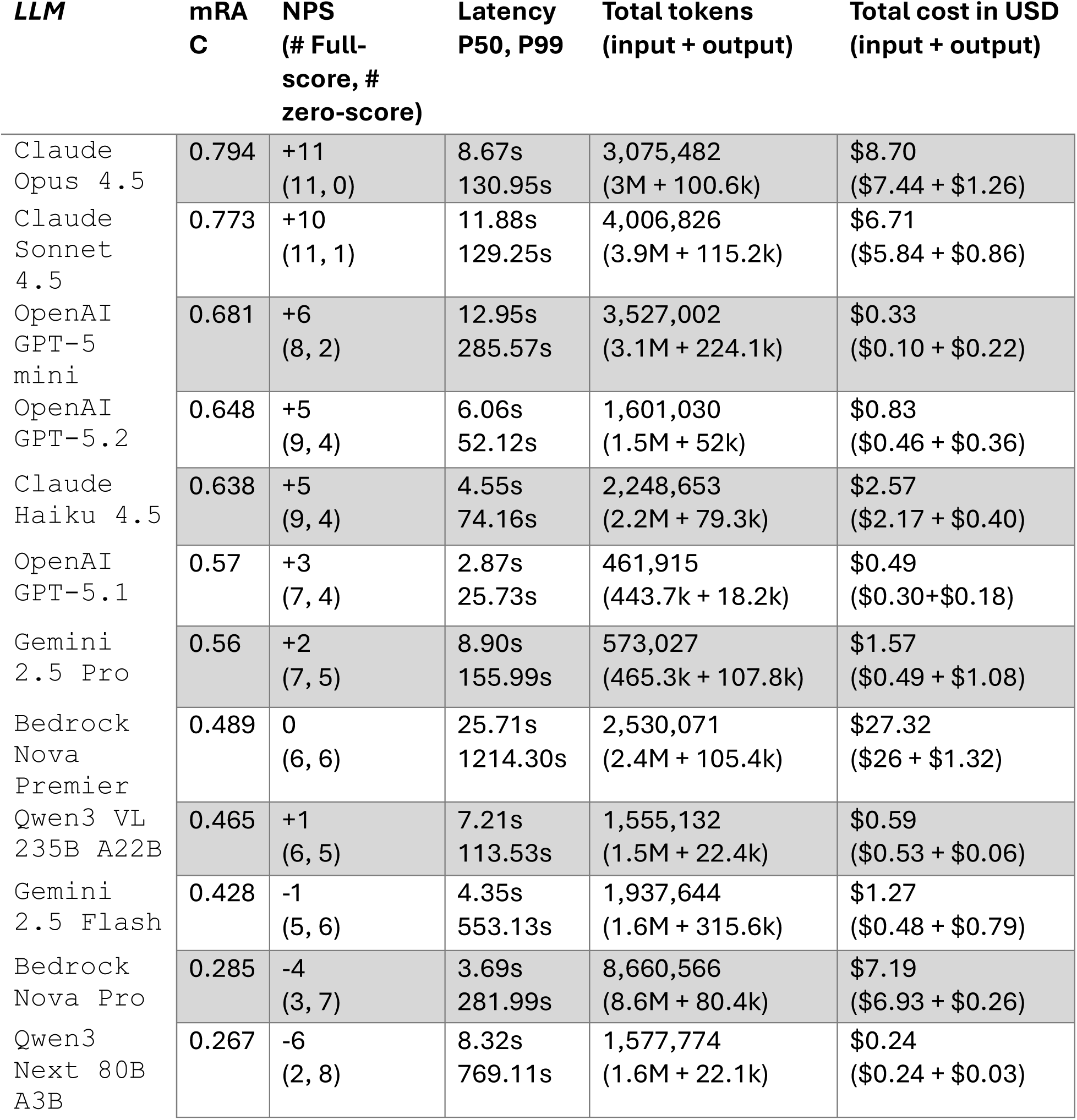
Detailed performance of various LLMs on the numeric response benchmark. . Cost and latency metrics are obtained from LangSmith tracing. The `Preview 09 2025` version was used for `Gemini 2.5 Flash` as it was better performing over its subsequent official version. LLM, large language model; mRAC, arithmetic mean of the reasoning-accuracy composite score across CQTS; P50, 50^th^ percentile; P99, 99^th^ percentile.

**Figure 3:**
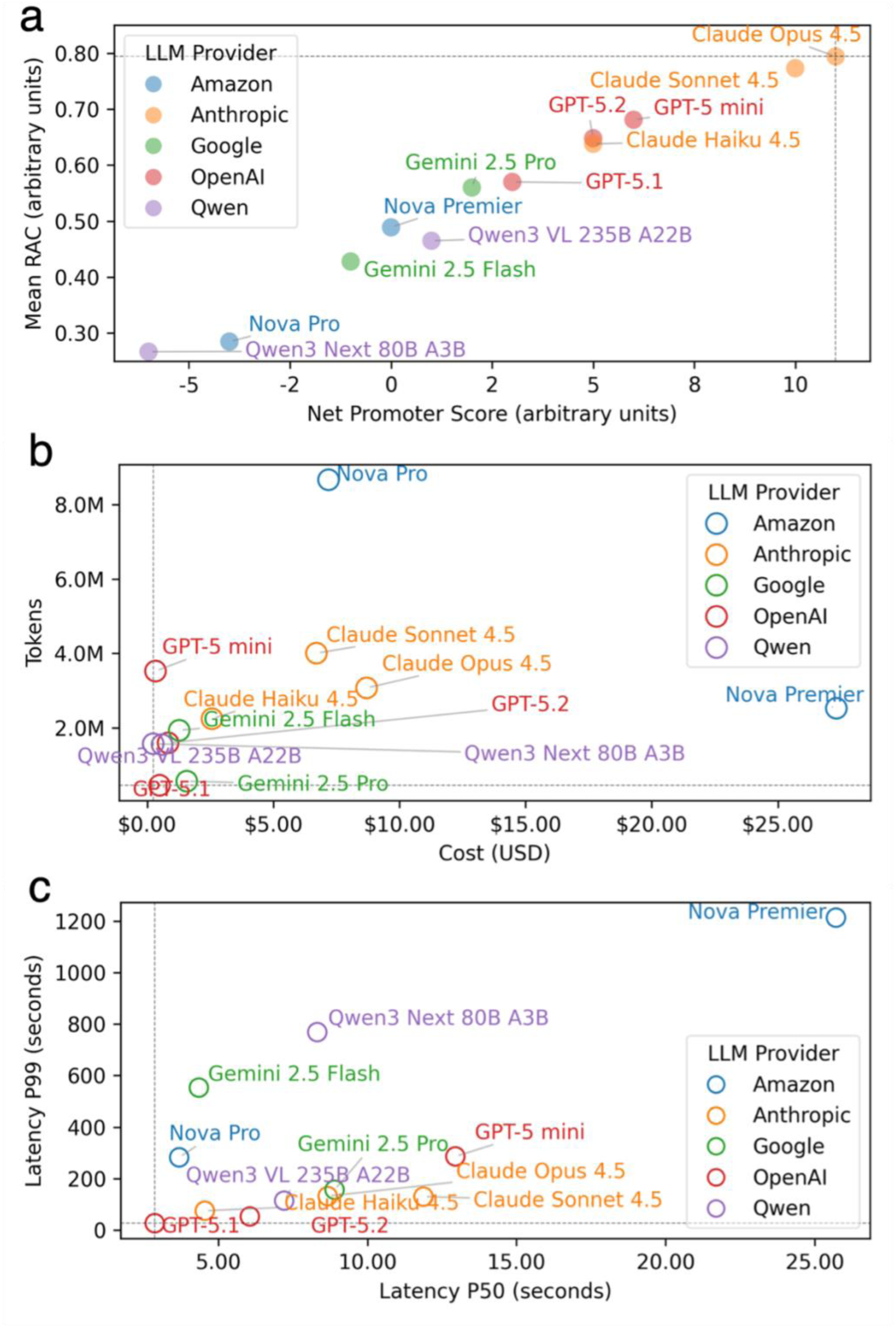
Numeric response benchmark performance of various LLMs. Performance metrics are grouped into (a) quality, (b) efficiency, and (c) latency. Dotted lines denote the best-in-class performance for that metric.

We compared LLMs based on their cost efficiency. While Qwen models were the cheapest to run the numeric response benchmark on, their useability is limited given their poor performance on mRAC and NPS (Figure 3a-b). The GPT-5 models from OpenAI were the next cheapest family of models, with token consumption below $1.00 per evaluation for all models (Table 4 rows 3, 4 & 6). GPT-5 mini was the most fastidious model with 3.5M tokens consumed, compared to 1.6M and 0.46M for GPT-5.2 and GPT-5.1. Google’s Gemini 2.5 Pro and Gemini 2.5 Flash models took the middle ground in both performance and cost, while Anthropic’s Claude 4.5 models were on the costly end. AWS Bedrock’s Nova Premier placed last in quality, price and latency (Figure 3c), while the lower-tier model Nova Pro performed poorly on quality metrics despite its high cost.

Taken together, the LLM choices can be narrowed down to three optimal configurations. Anthropic’s `Claude Opus 4.5` is recommended for maximum performance, provided latency and cost are not a concern e.g., locally hosted, single-user instances dedicated to computationally intensive tasks. If low latency and fast iteration is the priority, such as for simple repetitive tasks, OpenAI’s `GPT-5.1` model is preferred due to its brevity and speediness (medium latency 2.87s, 99^th^ percentile latency 25.73s, Table 4). For a balance between performance and cost, such as for deployment-at-scale and general tasks, OpenAI’s `GPT-5-mini` is our recommendation, being the least costly to evaluate yet ranking high on mRAC and NPS metrics (rank 3 out of 12, Table 4).

### Applying MyeGPT to known findings

We tasked MyeGPT to recapitulate known findings as a form of external positive control. This consists of two tasks with known results – 1. classification of functional high risk patients^23^, and 2. reproducing the PR vs non-PR RNA-seq subtype analysis^2^.

#### Reproducing Chung’s FHR analysis

Functional high risk (FHR) is an area of growing interest due to its aggressive clinical phenotype in the lack of known genomic risk markers. The ability of MyeGPT to classify FHR is of interest due to the need for various data modalities spanning cytogenetics, somatic mutations, CNAs, and clinical data. We tasked MyeGPT to reproduce a recent work by our lab^23^ on the analysis of risk factors contributing to FHR. We first manually classified 1143 NDMM into 250 GHR, 114 FHR, and 781 SR cases (heatmap in Figure 4a). The 781 SR cases was inclusive of 285 indeterminate cases, which had missing data in one or more modalities. We then tasked MyeGPT loaded with different LLMs to classify FHR and compared their results to those of the authors (patient level calls in Extended results 1).

When we calculated inter-rater reliability, the highest agreement between any two test-takers was `Claude Sonnet 4.5` vs. human (κ = 0.965, Figure 4b), followed by human vs. `GPT-5.2` (κ = 0.952) The highest agreement between any two LLMs was observed between GPT-5 mini vs. GPT-5.2 (κ = 0.947), closely followed by `Claude Opus 4.5` vs. `GPT-5.2` (κ = 0.946). In terms of central tendency, the model which had highest agreement with *all other* models was OpenAI’s `GPT-5.2` (mean κ = 0.941, values from rightmost column of heatmap, Figure 4b), followed by the human rater (mean κ = 0.938). The model with lowest one-vs-rest agreement was `Claude Opus 4.5` (mean κ = 0.909).

**Figure 4:**
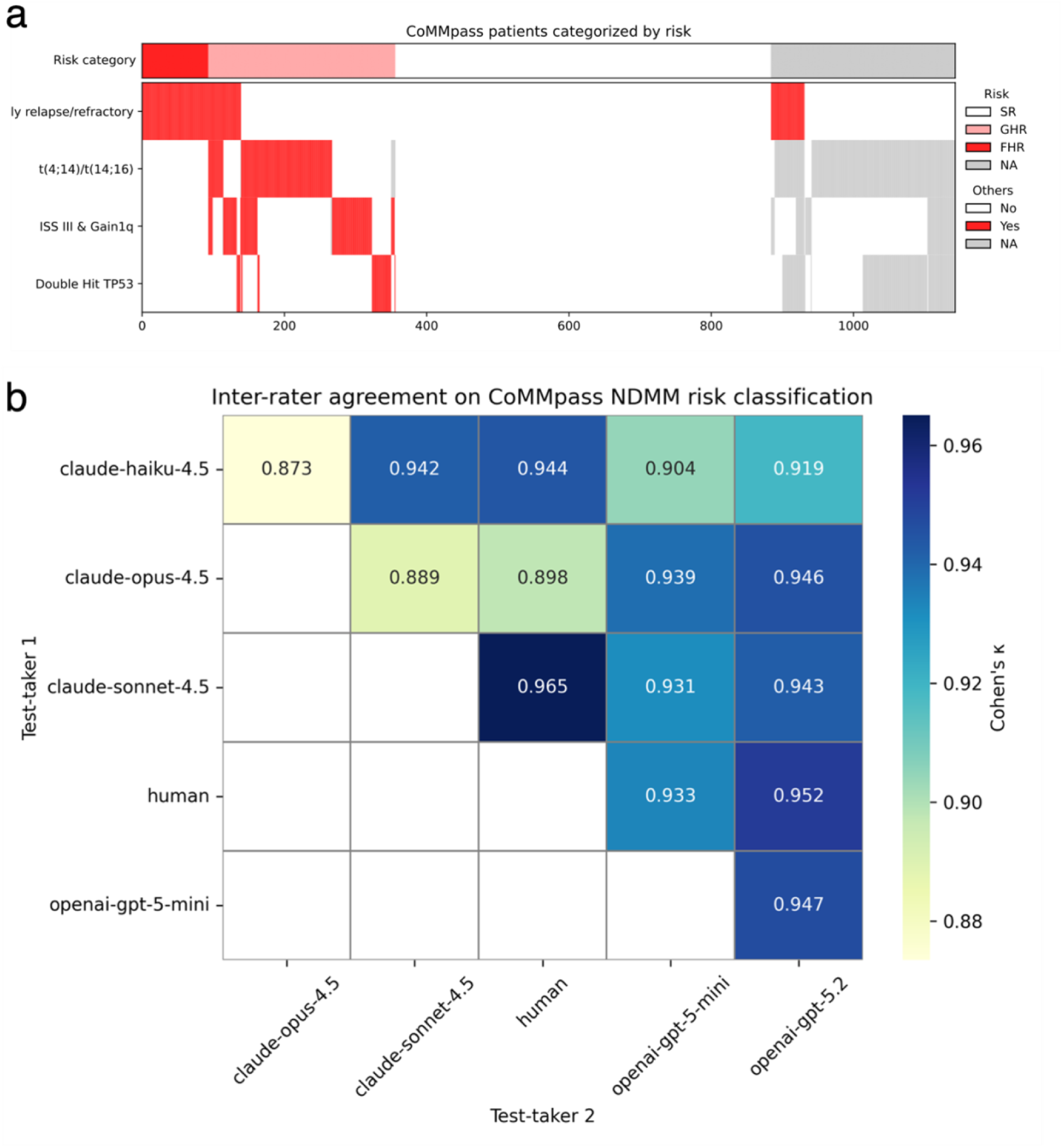
Reproducing FHR classification pipeline of Chung et al. (Submitted for publication). (a) NDMM risk classification into SR, GHR, and FHR. Heatmap shows the composing features, and top row shows the risk call. The 1143 NDMM patients were manually classified by J.G.C. with Python. (b) Inter-rater reliability between 5 LLMs and human on risk classification. Rather than relying on human as ground-truth, we opted for the unbiased approach of inter-rater reliability. A Cohen’s κ close to 1 indicates agreement between two classifiers. NDMM, newly diagnosed multiple myeloma.

When considering bottom-line agreement in the numbers per risk category, the highest concordance was observed in the high-risk translocation task (Table 5, row 3), where all test-takers were in complete agreement. This reflected the simplicity of the task compared to other tasks, since no merging of tables was required. The highest variability occurred in counting the Gain1q cases (mean absolute distance *M*. *A*. *D*. = 49.67) followed by counting the “refractory” subtype of clinical high-risk cases, where patients relapse after induction therapy or receipt of ASCT (*M*. *A*. *D*. = 36.27).

**Table 5.** NDMM risk classification bottom line responses. . The task was broken down into 5 steps. For the first task of identifying clinical high risk, “no response” is with reference to induction therapy, and “early relapse” is a progression within 12 months of induction therapy or ASCT. The LLMs evaluated here were the top five LLMs from the single-turn benchmark. For reference, responses from the author are included as “human”. ASCT, autologous stem cell transplant; FISH = fluorescent in-situ hybridization; FHR, functional high risk; GHR, genomic high risk; JGC, Jia Geng Chang; THC, Tae-Hoon Chung, LoH, Loss-of-heterozygosity; NS = non-synonymous; SR, standard risk.

| **Test taker** | **Human (JGC)** | **Human (THC)** | **Claude 4.5 Opus** | **Claude 4.5 Sonnet** | **Claude Haiku** | **GPT-5 Mini** | **GPT-5.2** |
| --- | --- | --- | --- | --- | --- | --- | --- |
| *Number of clinical high-risk cases* |  |  |  |  |  |  |  |
| 1. no response | 57 | 50 | 57 | 57 | 47 | 57 | 57 |
| 2. early relapse | 117 | 191 | 201 | 116 | 133 | 145 | 146 |
| 3. 1 OR 2 | 154 | 241 | 234 | 152 | 169 | 179 | 182 |
| *Number of cases of TP53 events* |  |  |  |  |  |  |  |
| 1. TP53 mutation | 56 | 54 | 56 | 48 | 56 | 56 | 56 |
| 2. TP53 Deletion | 65 | 42 | 79 | 65 | 79 | 79 | 79 |
| 3. TP53 LoH | 105 | <NA> | 104 | 86 | 105 | 104 | 105 |
| 4. 1 AND (2 or 3) | 36 | 15 | 36 | 30 | 36 | 36 | 36 |
| *Number of stage 3 cancer and Gain1q cases* |  |  |  |  |  |  |  |
| 1. ISS III | 311 | 311 | 311 | 218 | 311 | 311 | 311 |
| 2. Gain1q | 294 | 302 | 327 | 201 | 317 | 327 | 327 |
| 3. 1 AND 2 | 93 | 102 | 106 | 93 | 97 | 106 | 106 |
| *Number of high-risk translocation cases* |  |  |  |  |  |  |  |
| 1. t(4;14) | 116 | <NA> | 116 | 116 | 116 | 116 | 116 |
| 2. t(14;16) | 37 | <NA> | 37 | 37 | 37 | 37 | 37 |
| 3. 1 OR 2 | 152 | 169 | 152 | 152 | 152 | 152 | 152 |
| *Final task: risk categorization* |  |  |  |  |  |  |  |
| 1. GHR | 250 | 253 | 260 | 254 | 254 | 260 | 260 |
| 2. FHR | 114 | 136 | 159 | 115 | 126 | 123 | 127 |
| 3. SR | 781 | 754 | 724 | 785 | 765 | 762 | 758 |

#### Reproducing Skerget’s PR subtype analysis

As a positive control, MyeGPT was tasked to recapitulate published findings. We prioritized highly cited, external publications that used data from CoMMpass IA22. We narrowed down to the work of Skerget et al 2024, Nature Genetics^2^, the official publication accompanying the release of IA22. Of interest was Figure 3c, 3d, and 3e, involving survival analysis and differential gene expression on the Proliferative (PR) RNA-seq subtype of MM. We tasked MyeGPT to reproduce the calculations and plots as close to the reference as possible and assessed how close were the effect sizes and p-values.

First, MyeGPT was tasked to compare OS between PR and non-PR patients (Figure 5a). MyeGPT reported the median OS for PR to be 21.0 months (95% CI 14.8-54.5) versus 21.3 months (95% CI 15.0-55.3) in the reference publication. Median OS for Non-PR was found to be 102.5 months (95% CI 96.1-NA) for MyeGPT versus the reference 103.9 months (95% CI 97.4-NA). The computed log-rank p value of 1.032x10^-10^ almost matches the p-value of 1.1x10^-10^ of our reference publication. Upon investigation, we found the discrepancies in KM curves to originate from five patients whose right-censored overall survival differed between MyeGPT’s source files and those used in the publication (details in Extended results 2).

**Figure 5:**
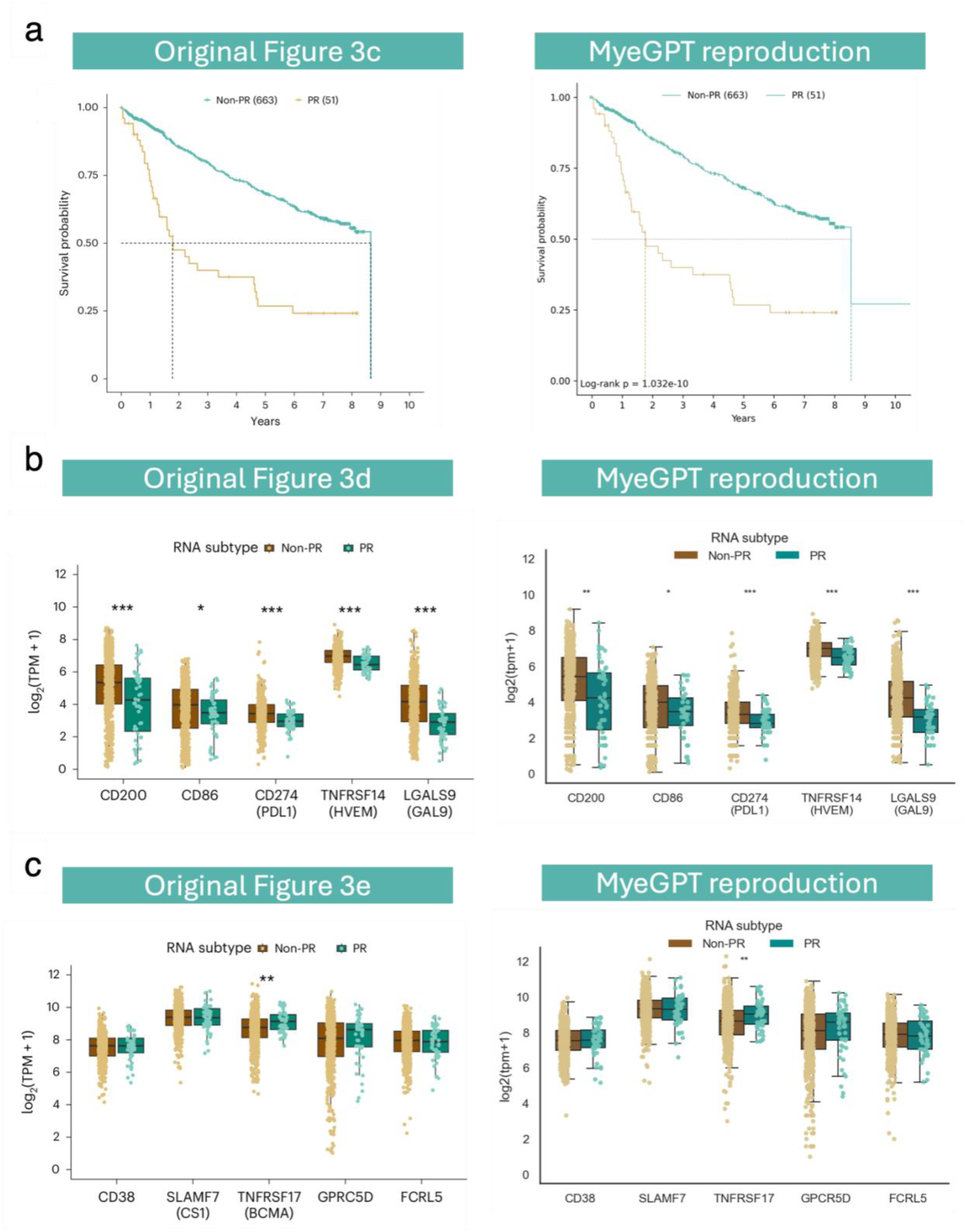
Reproducing the PR subtype analysis of Skerget et al 2024. We instructed MyeGPT using hand-crafted prompts to recreate three analyses related to the RNA-seq PR subtype from Skerget et al. 2024^2^. Raw values are in <u>Extended results 2</u>. (a) Kaplan-Meier analysis of overall survival between PR and non-PR. (b) Expression differences in checkpoint inhibitor genes. (c) Expression differences in immune genes. Asterisks above the box-and-whisker plots denote significance in Mann-Whitney U test between PR and non-PR subpopulations: (*) p<0.05; (**) p<0.01; (***) p<0.001.

We next tasked MyeGPT to compare the expression of five checkpoint inhibitor target genes *CD200*, *CD86*, *PD-L1*, *HVEM*, and *GAL9* between PR and Non-PR (Figure 5b), and similarly for five immune genes *CD38*, *SLAMF7*, *TNFRSF17*, *GPRC5D*, *FCRL5* (Figure 5c). Across the ten genes, the effect sizes and p-values of MyeGPT’s Mann-Whitney U tests largely matched the publication when comparing by significance levels. The only exception was the checkpoint inhibitor target *CD200*, for which MyeGPT obtained p-value of 0.00105 when it was p<0.001 in the reference publication. We suspect differences could be due to differences in batch correction, which we used `pyComBat`^21^ whereas the reference publication uses the `sva` R package. These small differences in batch corrected-tpm values also led to slight inconsistencies in MyeGPT’s re-constructed Bergsagel index values (Figure 6; raw values in Extended results 3), although the impact on test statistics were minimal.

**Figure 6:**
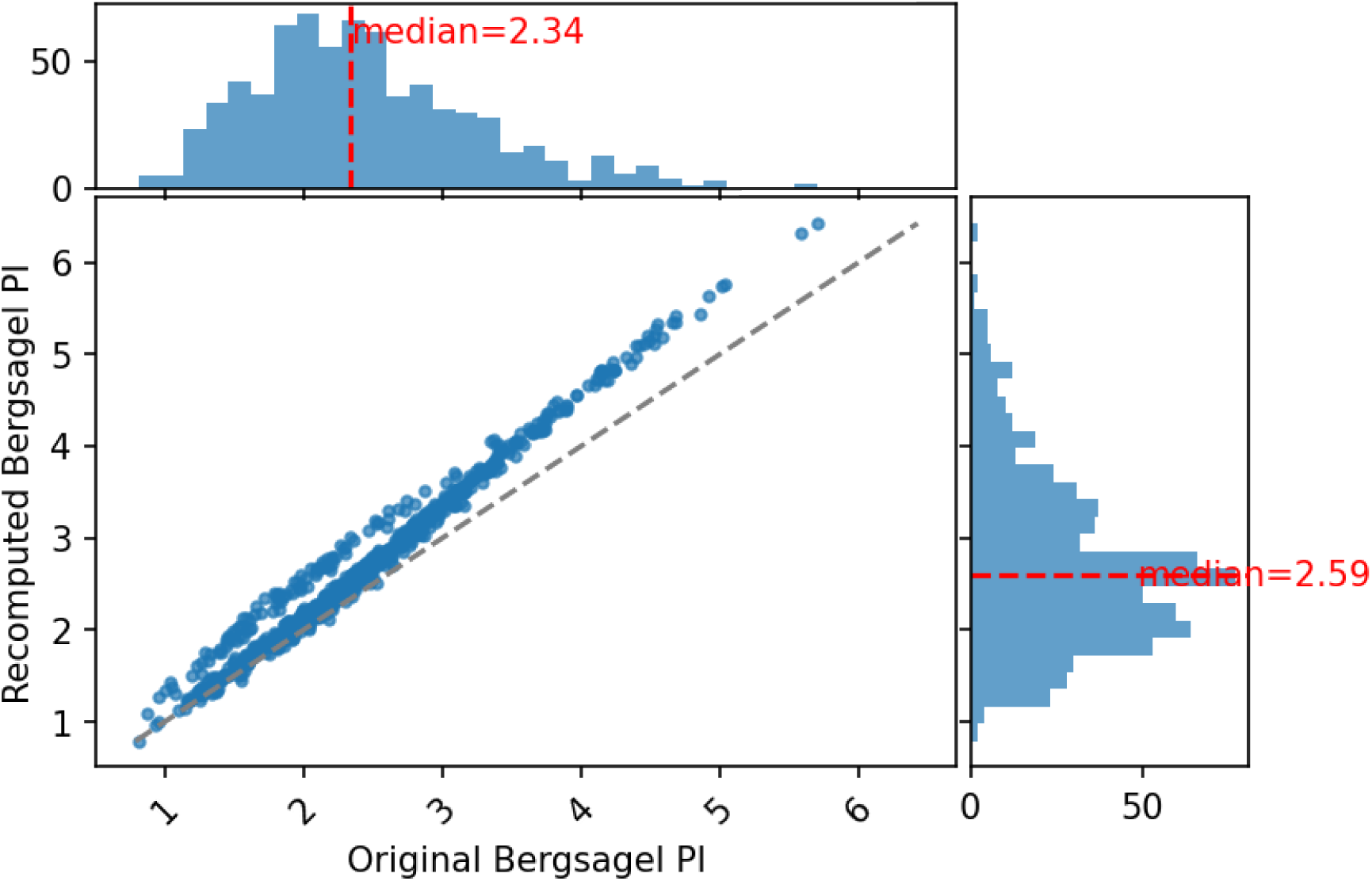
Reproduction of the Bergsagel Proliferation Index in Skerget et al. 2024^2^. Comparison of original versus reproduced PI values. Marginal histograms are shown. Dashed line indicates identity. We asked MyeGPT to calculate the Bergsagel proliferation index as the arithmetic mean log2(tpm + 1) expression of 12 genes: *TYMS, TK1, CCNB1, MKI67, PCLAF (KIAA0101,* ENSG00000166803*), GINS1 (KIAA0186,* ENSG00000101003*), CKS1B, TOP2A, UBE2C, ZWINT, TRIP13* and *KIF11.* Gradient > 1 is observed possibly due our pseudo-count choice of 1 being larger than Skerget et al. 2024, and variation about the skew could be due to differences in implementation of ComBat batch correction algorithm between PyComBat (us) and ComBatR (reference publication). Among N=712 samples, the difference is 0.285 ± 0.196 (mean ± standard deviation; recomputed PI is higher).

### Applying MyeGPT to novel scenarios

We demonstrate the type of analyses suited for MyeGPT by prompting it with a range of tasks with variable complexity. Tasks were arbitrarily classified into three tiers of complexity: *simple*, *intermediate*, and *complex*. We designed a total of eight tasks – two *simple*, four *intermediate*, and two *complex* tasks for which we had manual solutions beforehand (see Supplementary Text 3).

#### *Simple* tasks are single table univariate queries

Low complexity queries involve a single variable from one table in the CoMMpass database – of the form “what is the distribution of variable X”? For example, visualising the most frequently administered first line treatments (Figure 7a) and the top reasons for terminating first-line treatment (Figure 7b). Other queries in this tier can be to “classify patients into high and low expression of gene *G*”, “obtain the quantiles of blood LDH levels”, and “Find how many patients have mutations in gene *G*”.

**Figure 7:**
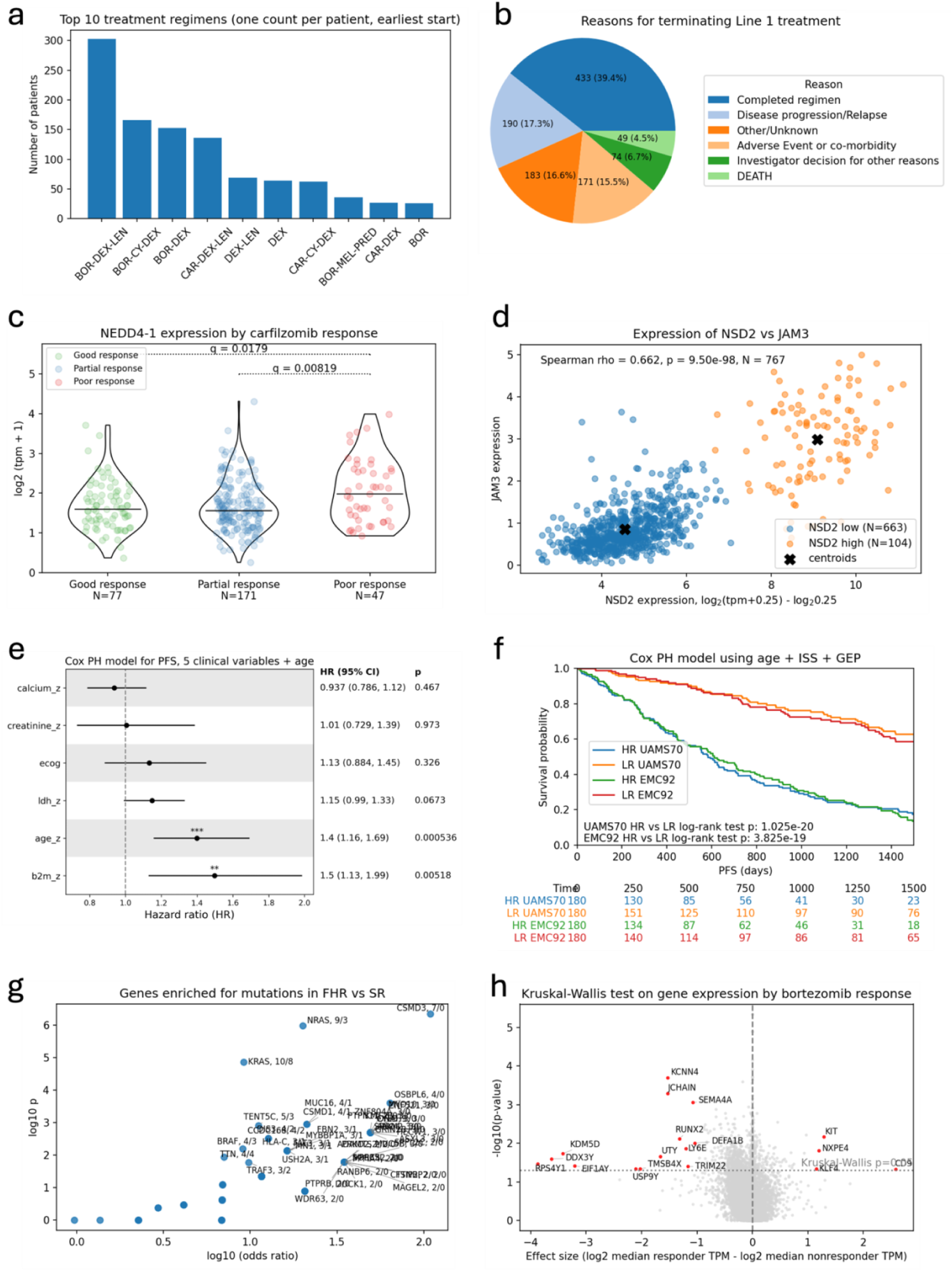
Applying MyeGPT on a range of scenarios. We put MyeGPT through 8 tasks with varying levels of complexity. (a) & (b) Low complexity queries involve single variable analysis; (a) frequency of top ten co-administered drugs as part of first line therapy. (b) Top five reasons for terminating first line therapy. (c),(d),(e),(f) medium complexity queries are the most common type of queries. They require the agent to merge data across variables and/or perform hypothesis testing or regression. (c) Differences in expression of *NEDD4-1* between good-, partial-, and non-responders of carfilzomib treatment. (d) K-means clustering of plasma cells by *NSD2*-*JAM3* expression profile. (e) Cox PH summary statistics of a clinical model for PFS. (f) KM survival curves of high- and low-risk patients predicted by age, ISS, UAMS70, and EMC92. (g), (h) High complexity analyses involve comprehensive statistical testing across feature sets like gene expression or variant calls. (g) Fisher’s exact test for genes enriched for mutations in functional high risk MM cases. (h) genes with unadjusted p<0.05 and effect size>1 are coloured red and annotated using gene symbol.

#### *Intermediate* tasks involve multivariate analysis

Medium complexity queries are hypotheses-driven question in the form of “what is the pattern between variables X, Y, and Z?”. They involve two or more variables, possibly from different clinical/omics modalities. Tables must be merged using INNER JOIN operations on columns such as gene ID or patient identifier. Any query involving statistical analyses e.g., likelihood-ratio tests, computing summary statistics, regression also fall into this category. As we expect most queries to be hypothesis-driven in nature, medium complexity queries fit the typical use case of MyeGPT. For example, comparing expression of *NEDD4-1* by response to carfilzomib (Figure 7c), K-means clustering of CD138-posiitve plasma cells by *NSD2*-*JAM3* expression profiles (Figure 7d), a Cox Proportional Hazards model of clinical variables (Figure 7e; raw values in Extended results 4), and survival analysis of patients stratified by UAMS70/EMC92 risk classification (Figure 7f; raw values in Extended results 5).

#### *Complex* tasks involve genome-wide screens

The most computationally expensive family of queries are non-hypothesis driven and part of explorative screens that involve extensive searching for statistically significant relationships (“find everything associated with X”). Queries that require data for all genes’ expression across all patients also fall under this tier, as it involves over 40 million data points of the PostgreSQL table for gene expression.

In Figure 7g, MyeGPT was tasked to identify genes enriched for mutations in FHR patients compared to standard risk patients (SR). To alleviate the burden of reclassifying NDMM cases into FHR and SR, the labels were supplied by the user using the outputs of Reproducing Chung’s FHR analysis. At the adjusted significance level of 0.05, 44 genes were significantly enriched in FHR (FDR q<0.05, Extended results 6). The known driver activity of majority of the genes significantly enriched in FHR patients (*NRAS* (rank 2), *KRAS* (rank 3), *TENT5C* (rank 7)), cancer biomarkers (*MUC16* (rank 5), *MUC5B* (rank 9), *MYO10* (rank 10)), and tumour suppressors (*CSMD3* (rank 1), *CSMD1* (rank 6)) suggests biological significance of these mutations in contributing to early relapse and drug refractoriness seen in FHR.

In Figure 7h, we tasked MyeGPT to find genes associated with responsiveness to bortezomib. MyeGPT compared the expression of 19850 genes between N=29 non-responders (disease remained stable or progressed after bortezomib-based treatment) and N=477 responders. Top results included cell volume regulator *KCNN4* (Η=13.8, p=0.0002, N_A_=504, N_B_=24), inhibitor of inhibitor *VEGF*-mediated angiogenesis *SEMA4A* (H=11.1, p=0.0009) and regulator of osteoblast differentiation *RUNX2* (H=7.14, p=0.007; Extended results 7).

Other examples of high-complexity queries include performing gene-wise Cox regression across all protein coding genes to find survival-relevant genes, and gene-wise correlation analysis to find co-regulated genes with the gene of interest.

## Discussion

A major innovation of MyeGPT lies in the integration of agentic-AI into multiple myeloma analysis. We combined the CoMMpass database, our custom annotations, and a custom knowledge base into the ReAct framework – packaged into a web-based interface for user convenience. While there exists an LLM-powered chatbots for multiple myeloma (see *Myelo*, Supplementary Table 1), it is merely a chatbot for patients to answer frequently asked questions. In comparison, MyeGPT is the first AI agent tailored for multiple myeloma researchers. MyeGPT also demonstrates the proof-of-concept for the *dataset curator*—an agent which is highly knowledgeable about one dataset. Be it for multiple myeloma or other diseases, we envision a scenario where research groups create specialised *dataset agents* to converse with their datasets of interest.

Besides the concept of dataset agents, other aspects of this manuscript may be of use the biomedical agent space. One is with respect to cost optimisation. Evaluation of initial versions of MyeGPT was costing ∼$25 per run of CQTS. We narrowed down the issue to the mandatory parsing of the lengthy documentation as part of agent initialisation, which contributed greatly to input token usage. To overcome this, we notice that not all tables are required in answering typical questions. By paginating documents by data modality and employing RAG on user queries, only specific pages of documentation need to be retrieved, as and when it is needed. This avoids context decay, led to a slight drop in response latency, and greatly reduced token consumption.

Another token-friendly technique we employed was to leverage “code-actions” and discourage “text-actions ^4^”. In the initial versions of MyeGPT, the ‘`LangChain SQL query`’ tool was generating >1M tokens per step as it returns query results as printed text. To overcome this, we enforced LIMIT 3 clauses to all queries written by this tool, returning a result-snippet just sufficient for debugging. Once a correct SQL query is built, the actual SQL query is executed by Python and the (potentially large) results are silently written to disk. By combining RAG-as-a-tool and minimizing text-actions, among other optimisations, we reduced the cost per run of the numeric response benchmark by 100-fold, or from $10 to $0.10 for OpenAI’s GPT-5 mini.

As a new tool at the time of writing, a few limitations of MyeGPT are worth noting. The main limitation is its restriction to research environments. The non-deterministic nature of LLMs mean that clinical decisions solicited or emitted from the software ought to be taken lightly or disregarded. Moreover, in the ideal scenario where MyeGPT was always accurate, the reliability of its clinical recommendations is still low compared to double-blind case-control trials, due to the observational and open-label nature of the CoMMpass study. Even so, in research-based scenarios where specificity is prioritised over sensitivity i.e. relapse prediction vis-à-vis biomarker discovery, the onus is on the researcher to re-run the analysis using the same set of input data. Together, the probabilistic nature of its underlying LLMs and its limited accuracy restrict the applicability of MyeGPT to research settings i.e. exploratory data analysis and hypothesis generation.

Another limitation is the concern over patient data sensitivity. Unless the LLM is hosted locally, patient data is unavoidably passed between the servers of LLM providers. While this is less of an issue for publicly available, de-identified datasets like CoMMpass—where patients identities and germline variants are masked—AI agents for in-house or private datasets may need to self-host LLMs entirely on-premises. In addition, they must implement data safety guardrails to prevent database-destruction attacks or the export of sensitive data through malicious prompts. With agentic AI, extra scrutiny over the egress of patient data is required, whose ability to write and execute code confers it *carte blanche* over its actions.

## Supporting information

Extended Results

## Acknowledgements

We thank the organizers and participants of the CoMMpass study, the Multiple Myeloma Research Foundation for sponsoring the study, Ms. Baohong Lin and Mr. Tze King Tan for their assistance in cloud computing and software credits acquisition, Dr. Irfan Azaman and Dr. Reinhard Brunmeier from Cancer Science Institute, Singapore and Dr Hearn Jay-Cho from the MMRF for their feedback on the software, and Dr Joan Sim Poh Ling and Mr Jonathan Tan from NUS Technology Transfer and Innovation Office for licensing advice.

## Data availability

MyeGPT’s source code is accessible at https://github.com/JiaGengChang/MyeGPT. It is open source for non-commercial use under the GNU General Public License V3. The 20-question CoMMpass quantitative test set is deposited at https://smith.langchain.com/o/5b1493fe-9a9d-483f-aa31-5ce007110424 as a public eval dataset.

## Author Contributions

J.G.C. was responsible for the conceptualisation and development of the bioinformatic tool, creating the evaluation benchmarks, and writing the manuscript.

A.M.G. was responsible for the integration of MyeGPT into the MMRF Virtual Lab analysis suite, analysing the experimental results, and the editing of the manuscript.

J.R. was responsible for integrating the software into the MMRF Virtual Lab analysis suite.

T.H.C. was responsible for testing the multi-turn prompting benchmark.

W.J.C. and G.M. were responsible for guiding the direction of the study, analysing the experimental results, and reviewing the manuscript.

## Clinical Trial number

Not Applicable

## Appendix

### Index of Extended Results

Extended results 1 **Risk classification bottom line responses**. Individual patient level classification into standard risk (SR), genomic high-risk (GHR), and functional high-risk (FHR) by two human test takers and five LLMs as part of Reproducing Chung’s FHR analysis.

Extended results 2 **Skerget PR re-analysis results**. The tabular results for Reproducing Skerget’s PR subtype analysis, including discrepancies in survival data of 5 non-PR patients between our version of IA22 clinical data and that used by Skerget et al. Nature Genetics 2024. The exact p-values, test statistic values, and confidence intervals reported here.

Extended results 3 **MyeGPT-reconstructed Bergsagel’s Proliferation Index in Figure 6**. Only first visit samples were used. Offset 1 denotes tpm+1 transformation before log transformation.

Extended results 4 **Clinical Cox PH model in Figure 7e**. Summary statistics of multivariate Cox Proportional Hazards model with clinical variables

Extended results 5 **Raw data for GEP KM curves of Figure 7f**. Only samples with uams_pred_group in LR or HR are shown in the curves. Same for emc_pred_group. Haz_ is hazard ratio. event 1 means progression; time denotes progression-free survival.

Extended results 6 **Fischer’s Chi-squared test for enrichment in FHR vs SR in Figure 7** Figure 6**g**. Only genes with ≥1 carriers in FHR are considered.

Extended results 7 **Results of Kruskal Wallis test for response to bortezomib, seen in Figure 7h**. The ‘responder’ and ‘nonresponder’ columns contain medium log2(tpm+1) values of either the responders or non-responders. Common across all tests: number of bortezomib responders (partial response minimum): 504. Number of non-responders: 24 (stable or progressive disease).

### Index of Supplementary Tables

Supplementary Table 1 **A survey of existing software developed for the CoMMpass study**. The MMRF Researcher Gateway was deprecated in September 2025 and is in the process of being revamped into the MMRF VirtualLab. Abbreviations: NCI-GDC, National Cancer Institute Genomic Data Commons; TCGA, The Cancer Genome Atlas; KM, Kaplan-Meier analysis; MMRF, multiple myeloma research foundation. Superscripts denote references.

Supplementary Table 2 **A list of data analysis agents in the biomedical space**. Examples of queries suited for these agents are included. These suggested workflows were obtained from their official websites.

Supplementary Table 3 **Overview of MyeGPT’s knowledge base**. File names and descriptions of the 52-page CoMMpass documentation used for knowledge retrieval. Each page stores conceptually related fields originating from the same table. While small tables such as canonical_ig and chromothripsis only have one document each, large tables per_patient and per_visit have 7 and 20 pages of documentation.

Supplementary Table 4 **The document retrieval dataset for the Knowledge retrieval benchmark**. Consists of mappings between 250 simulated queries to the intended reference document in the CoMMpass knowledge base.

### Index of Supplementary Texts

Supplementary Text 1 **The LLM-as-a-judge prompt for automated evaluation.** This script is the system prompt used for LLM-as-a-judge in the Numeric response benchmark. This prompt instructs the LLM to evaluate MyeGPT-generated answer against the authors’ model answers and assign a score based on the allocation rules for that question. Terms enclosed in curly braces – {inputs}, {outputs}, {reference_outputs} – are placeholders that will be dynamically substituted depending on the question.

Supplementary Text 2 **Prompts for Skerget’s PR subtype analysis** The prompts used to recreate figures 3c, 3d, and 3e of Skerget S. et al., *Nature Genetics*, 2024, producing the graphical results seen in Reproducing Skerget’s PR subtype analysis. The prompt for calculating the Bergsagel proliferation index (Figure 6) is also included.

Supplementary Text 3 **Prompts for use case demonstration**. This file contains the prompts used in Applying MyeGPT to novel scenarios. Each prompt contains the high-level task paired with low-level instructions, such as refinements to the plot title, colour palette, or whether to include p-value annotations.

Supplementary Text 4 **MyeGPT’s LLM system prompt.** The system prompt used for initialization of the LLM inside MyeGPT. The structure of the prompt follows an adaptation of the AIM framework: Ask with Purpose, Include Details, Modify and Refine. It frames the LLM as a data analyst for multiple myeloma and provides guidelines on tool use, problem-solving strategy, and stylistic guidelines

### Index of Supplementary Files

Supplementary File 1 **question-answer.ipynb** Manually typed Python code created to obtain ground truth answers for 13 of the 20 questions in the CoMMpass quantitative test set, used in the Numeric response benchmark.

Supplementary File 2 **call-fhr.ipynb** A script that author J.G.C. wrote for Reproducing Chung’s FHR analysis, to manually classify 1143 NDMM patients in CoMMpass into standard risk, genomic high risk, and functional high risk. This also contains 7 questions and answers as part of the 20-question CQTS.

## Footnotes

1 Pgvector: an open-source Postgres extension for similarity search

2 Psycopg: an open-source PostgreSQL driver for Python

3 REPL: Read, Evaluate, Print, Loop

4 Text-actions refer to actions which return printed text into the standard output. They cost output tokens as they are part of the LLM response. On the other hand, code-actions execute the SQL query in a Python environment, and the output to be silently written to disk. As a result, code-actions primarily only incur thinking tokens.

## References

1. Keats, J. J. et al. Interim Analysis Of The Mmrf Commpass Trial, a Longitudinal Study In Multiple Myeloma Relating Clinical Outcomes To Genomic and Immunophenotypic Profiles. Blood 122, 532 (2013).

2. Skerget, S. et al. Comprehensive molecular profiling of multiple myeloma identifies refined copy number and expression subtypes. Nat Genet 56, 1878–1889 (2024).

3. Fiala, M. A. et al. Variations in Multiple Myeloma Disease Presentation By Race. Blood 126, 5618 (2015).

4. Sharma, N. et al. The Prognostic Role of MYC Structural Variants Identified by NGS and FISH in Multiple Myeloma. Clin Cancer Res 27, 5430–5439 (2021).

5. Miller, C. et al. A Comparison of Clinical FISH and Sequencing Based FISH Estimates in Multiple Myeloma: An Mmrf Commpass Analysis. Blood 128, 374 (2016).

6. Palumbo, A. et al. Revised International Staging System for Multiple Myeloma: A Report From International Myeloma Working Group. J Clin Oncol 33, 2863–2869 (2015).

7. Goldsmith, S. R. et al. Next Generation Sequencing-based Validation of the Revised International Staging System for Multiple Myeloma: An Analysis of the MMRF CoMMpass Study. *Clinical Lymphoma*, Myeloma and Leukemia 19, 285–289 (2019).

8. Settino, M. & Cannataro, M. MMRFBiolinks: an R-package for integrating and analyzing MMRF-CoMMpass data. Brief Bioinform 22, bbab050 (2021).

9. Settino, M. & Cannataro, M. MMRFVariant: Prioritizing variants in Multiple Myeloma. Informatics in Medicine Unlocked 39, 101271 (2023).

10. Four AI Agent Strategies That Improve GPT-4 and GPT-3.5 Performance. Four AI Agent Strategies That Improve GPT-4 and GPT-3.5 Performance https://www.deeplearning.ai/the-batch/how-agents-can-improve-llm-performance/ (2024).

11. Huang, K. et al. Biomni: A General-Purpose Biomedical AI Agent. 2025.05.30.656746 Preprint at 10.1101/2025.05.30.656746 (2025).

12. Zhang, M. et al. PromptBio: A Multi-Agent AI Platform for Bioinformatics Data Analysis. 2025.07.05.663295 Preprint at 10.1101/2025.07.05.663295 (2025).

13. Superbio.ai. https://app.superbio.ai/copilot.

14. Yang, E.-W. & Velazquez-Villarreal, E. AI-HOPE: an AI-driven conversational agent for enhanced clinical and genomic data integration in precision medicine research. Bioinformatics 41, btaf359 (2025).

15. Cortés-Ciriano, I. et al. Comprehensive analysis of chromothripsis in 2,658 human cancers using whole-genome sequencing. Nat Genet 52, 331–341 (2020).

16. Chang, J. G., Chen, J., Chew, G.-L. & Chng, W. J. MyeVAE: a multi-modal variational autoencoder for risk profiling of newly diagnosed multiple myeloma. *BMC Artif*. Intell. 1, 8 (2025).

17. Lewis, P., et al. Retrieval-Augmented Generation for Knowledge-Intensive NLP Tasks. Preprint at 10.48550/arXiv.2005.11401 (2021).

18. Koh, M. Y. et al. The ADAR1-regulated cytoplasmic dsRNA-sensing pathway is a novel mechanism of lenalidomide resistance in multiple myeloma. Blood 145, 1164–1181 (2025).

19. Soekojo, C. Y., Chung, T.-H., Furqan, M. S. & Chng, W. J. Genomic characterization of functional high-risk multiple myeloma patients. Blood Cancer J. 12, 1–9 (2022).

20. Pilcher, W. C. et al. A single-cell atlas characterizes dysregulation of the bone marrow immune microenvironment associated with outcomes in multiple myeloma. Nat Cancer 7, 224–246 (2026).

21. Behdenna, A. et al. pyComBat, a Python tool for batch effects correction in high-throughput molecular data using empirical Bayes methods. BMC Bioinformatics 24, 459 (2023).

22. Yao, S., et al. ReAct: Synergizing Reasoning and Acting in Language Models. Preprint at 10.48550/arXiv.2210.03629 (2023).

23. Chung, T.-H., Chang, J. G., Tan, T.-K. & Chng, W. J. Augmentation of the IMS/IMWG high risk criteria for prognosis in multiple myeloma. Haematologica (Submitted for publication).

