## Supplementary Texts for "“MyeGPT: an AI agent for Multiple Myeloma”"

Supplementary Text 1 **The LLM-as-a-judge prompt for automated evaluation.** This script is the system prompt used for LLM-as-a-judge in the Numeric response benchmark. This prompt instructs the LLM to evaluate MyeGPT-generated answer against the authors’ model answers and assign a score based on the allocation rules for that question. Terms enclosed in curly braces – {inputs}, {outputs}, {reference_outputs} – are placeholders that will be dynamically substituted depending on the question.

```
CORRECTNESS_PROMPT = """You are an expert data labeller evaluating model outputs for correctness.

<Instructions>
  - Carefully read the input and output
  - Check for factual accuracy
  - Focus on correctness of information rather than style or verbosity
</Instructions>

<Reminder>
  The goal is to evaluate factual correctness and completeness of the response.
</Reminder>

The input consists of the user's question (compulsory), definitions (optional), hints (optional):
<input>
{inputs}
</input>

The output consists of the answer (compulsory), scoring (optional), remarks (optional), and SQL code (if applicable):
<output>
{outputs}
</output>

Use the reference outputs below to help you evaluate the correctness of the response:

<reference_outputs>
{reference_outputs}
</reference_outputs>

Follow the scoring criteria provided in the input, and accept minor numerical deviations as specified:

<Tolerance>
Not all answers must be exact. Allow the following tolerances:
For answers <10, must be exact
For answers 10-50, correct if within +/-1
For answers 50-100, correct if within +/-2
For answers >100, correct if within +/-3
e.g. If the reference answer is 45, then 44, 45, or 46 are all acceptable.
</Tolerance>

"""
```

Supplementary Text 2 **Prompts for** **Skerget’s PR subtype analysis** The prompts used to recreate figures 3c, 3d, and 3e of Skerget S. et al., *Nature Genetics*, 2024, producing the graphical results seen in Reproducing Skerget’s PR subtype analysis. The prompt for calculating the Bergsagel proliferation index (Supplementary Figure 1) is also included.

### Figure 5a of main text / Figure 3c of reference publication

The 12 RNA subtypes of 714 patients are in result/skerget_patient_features.csv, with public ID stored under "Patient_ID", and RNA subtype stored under "RNA_Subtype_Name".

Perform kaplan meier survival analysis of PR vs non-PR patients and plot the survival curves.

Use teal color (#59b5a9) for Non-PR and gold for PR (#d2b172)

Add horizontal and vertical lines to indicate the median survival for PR and non-PR.

Trim the vertical lines at 0.50. Do not show them beyond y=0.50. trim the horizontal dotted line along y=0.50 to end at the non-PR median OS value

For x axis, convert the units from days to years. Use tick labels 0, 1, 2, ... 8, 9, 10. The x limit is to be -0.5 to 10.5

For y axis, use y limits -0.05 to 1.05, with tick marks and labels at y=0, 0.25, 0.5, 0.75, and 1

Name the x axis Years, y axis Survival probability. No plot title.

Show censored events (censor flag = 0) as vertical marks (|) on the curve. make these vertical bars smaller than default. e.g. markersize=4

remove the upper and right splines

Set legend to ncol=2 and border alpha 0, position top right. name the legend handles as Non-PR (558) and PR (43)

make the font size of the legend handles as large as the font size of the tick labels, and set the legend position to upper center

perform log rank test and get the p value between Non-PR and PR survival curves. Annotate on the bottom left corner of the plot the log rank p-value.

Save the figure as 300dpi, height 5.5in, width 6in, use a narrower line width for the two curves

Now report statistics like median OS with 95% confidence intervals, and the log rank test statistics

### Figure 5b of main text / Figure 3d of reference publication

Plot a pairwise boxplot with the 5 genes CD200, CD86, CD274 (PDL1), TNFRSF14 (HVEM), and LGALS9 (GAL9) along the x axis and Non-PR/PR subtype as position dodge, and log2(tpm+1) as the y axis.

Then plot pairwise jittered scatterplot over the boxplots, with the same x and y axes. Use a jitter value of 0.1.

Use CD200, CD86, "CD274\n(PDL1)", "TNFRSF14\n(HVEM)", and "LGALS9\n(GAL9)" as x axis labels, unrotated.

For the y axis, set the y limits to -0.5 to 12.5, and use tick labels at 0, 2, 4, 6, 8, 10, and 12

Use the following colors for the box (box) and scatter (pts):

non_pr_box = "#8c5a25" (dark brown)

pr_box = "#008b8b" (teal)

non_pr_pts = "#d9c289" (light tan)

pr_pts = "#6ec4b6" (light teal)

Between PR and Non-PR groups for each gene, perform a two-sided unpaired Wilcoxon rank sum test and are indicated when significant (*P < 0.05, **P < 0.01 and ***P < 0.001). if the result is non-significant, indicate using ns. Set the y-value of these text values to 10.

Remove the top and right splines

For legend, title the legend RNA subtype. Use two handles: Non-PR and PR. Use ncol=2, bbox to anchor x=0.5, y=1.12, legend border alpha 0, marker='s'

### Figure 5c of main text / Figure 3e of reference publication

Now repeat for the following genes:

CD38, SLAMF7 (CS1), TNFRSF17 (BCMA), GPCR5D (Gene stable ID is ENSG00000111291), FCLR5 (Gene stable ID is ENSG00000143297)

Use as the x-axis labels: "CD38", "SLAMF7\n(CS1)", "TNFRSF17\n(BCMA)", "GPCR5D", and "FCLR5"

### Figure 6 of main text (original vs recomputed Bergsagel’s PI)

Recompute Bergsagel PI using the geometric mean expression of the log2(TPM+)-transformed expression of the following 12 genes: TYMS, TK1, CCNB1, MKI67, PCLAF ( KIAA0101), GINS1 (KIAA0186), CKS1B, TOP2A, UBE2C, ZWINT, TRIP13 and KIF11.

Now plot a scatterplot of the Bergsagel PI that is recomputed, versus the values inside result/skerget_patient_features.csv under the column "Proliferation_Index_Bergsagel".

Supplementary Text 3 **Prompts for use case demonstration**. This file contains the prompts used in Applying MyeGPT to novel scenarios. Each prompt contains the high-level task paired with low-level instructions, such as refinements to the plot title, colour palette, or whether to include p-value annotations.

### Analysis 1 – for Figure 7a

Produce a pie chart of the reasons patients to terminate first line treatment, e.g. completed regimen, relapse, etc.

Do not double count patients. The total number of cases should sum to less than or equal to the N=1143 patients.

Classify minor events with frequency <4% as "Other/Unknown”.

Set plot title as "Reasons for terminating Line 1 treatment”.

Do not use plt.tight_layout()

### Analysis 2 – for Figure 7b

Find the top 10 frequently administered drugs as part of first-line therapy. Then produce a heatmap showing the counts of co-administration among these 10 drugs. Show only the upper right triangle and include the counts in the heatmap as text annotations.

Use treatment short names e.g. bor for bortezomib. Rotate both axes' tick labels by 45 degrees.

For colormap use 'Reds'

### Analysis 3 – for Figure 7c

Plot a scatterplot comparing log2tpm expression of NEDD4-1 by response to carfilzomib, using some jitter.

Classify patient's response to carfilzomib treatment as Good (vGPR, CR), Partial (PR), and Poor  (SD, PD).

Overlay violin plots onto the scatter plot, using just outline and no fill color. Include horizontal line for median log2tpm.

For colors use the first 3 colors of matplotlib cmat Set1, in the order Poor (cmap idx 0), Partial (cmap idx 1), Good (cmap idx 2)

Perform Kruskal wallis test between good vs poor and partial vs poor. Correct for multiple testing using Benjamini-Hochberg to get q-values. Add text annotation of q values (3sf) together with a horizontal q value bar to y-positions y=5 and y=5.2, x-positions being the x-values of good/partial/poor response.

Set the title to "NEDD4-1 expression by carfilzomib response"

Set the x-tick labels as Good/Partial/Poor response (N=xxx) where xxx is the number of patients in that category.

### Analysis 4 – for Figure 7d

I want to compare the correlation of my reference gene NSD2 expression with the query gene C19orf81 across all first visit RNA-seq samples. Perform log2 ({TPM} + 0.25) - log2(0.25) transform on the expression values, then record the Pearson correlation and p-value.

Plotting instructions:

Title: "Expression of NSD2 vs JAM3"

Axis labels: y=JAM3 expression, x = r'NSD2 expression, log$_{2}$(tpm+0.25) - log$_{2}$0.25'

Primary task: Scatterplot with alpha=0.5

Secondary task: Perform K-means clustering with K=2 and mark centroids with 'x'.

Fill color: Use the cluster labels to decide the fill color of the observations.

Legend position: bottom right

Legend handle names: "NSD2 low (N={N_low})", "NSD2 high (N={N_high})", "centroids"

Text annotations: "Pearson correlation = {R}, p = {p}, N = {N}"

### Analysis 5 – for Figure 7e

Perform multivariate Cox regression using the following clinical parameters: B2M levels, creatinine levels, LDH levels, age, and ECOG performance status. Perform z-score transformation for age and lab values.

Plot a forest plot to visualize the results of the Cox regression.

Order the variables by hazard ratio. Add a vertical dashed line around HR = 1. Use linear scale for the hazard ratio.

Use alternating grey and white background colors for each row in the plot. Set the y limit to [ -0.5*min_y_value * row_height, max_y_value + 0.5 * row_height ], where row_height is the difference between consecutive y-values.

Include HR (95% CI) and p value (3 sig figures) as external columns to the right of the plot. For HR, use the format HR (lower 95 CI, upper 95 CI). Set col gap to 0.5. Offset the x-start of the HR column by 0.05.

Do not plot using plt.tight_layout().

Set plot title to "Cox PH model for PFS, 5 clinical variables + age"

### Analysis 6 – for Figure 7f

Build 2 sets of Cox PH models for PFS. Model 1. age + ISS + UAMS70, Model 2. age + ISS + EMC92, where UAMS70 and EMC92 are scalar-valued GEP risk scores.

Predict the hazard using the fitted functions and plot the KM curves of the top 25% and bottom 25% of patients, overlaying the 4 curves.

Perform log-rank test on each pair of curves, between HR UAMS70 vs LR UAMS70 and between HR EMC92 and LR EMC92.

Plotting instructions:

Plot: 2 pairs of KM survival curves

Limits: x limit between 0 and 1500, y limit between 0 and 1

Legend: place the legend on 'left bottom' with a positive y offset of 0.2

Legend handle names: HR UAMS70 for top 25% of the UAMS70 model, and LR EMC92 for the bottom 25% model, etc. Deduce the names of the other two.

Title: Title the plot "Cox PH model using age + ISS + GEP"

Aspect ratio: Use 6x5 inches, 150 dpi

Text annotations: Annotate "UAMS70 HR vs LR log-rank test p: {p-value}" and "EMC92 HR vs LR log-rank test p: {p-value}" the bottom left of the plot.

Colors: For colors use the first 4 colors of cmap tab10.

Additional info: Show at risk counts

### Analysis 7 – for Figure 7g

The prompt is delivered in two stages. One to compute the results (row 1), another to plot the results (row 2).

Part 1 (analysis)

I have classified patients into functional high risk (result/human/functional_hr.csv) and standard risk (result/human/standard_risk.csv).

For one gene at a time, Perform fisher's exact test of FHR vs SR for the number of carriers vs non-carriers of the gene

Filter for genes:

non-immunoglobulin gene

non-Y chromosome gene

>2 events in FHR + SR combined

Save the test statistics to "fhr_vs_sr_fisher_no_igfilter_totalge3.csv"

Part 2 (plotting)

The fisher test statistics for differentially mutated genes between FHR vs SR are saved in "result/fhr_vs_sr_fisher_no_igfilter_totalge3.csv"

Take the rows of this file with non-NA gene symbol

Plot a volcano plot of the neglog10 p value against log10 odds ratio

Plotting instructions:

Plot: scatterplot, alpha 0.8

Title: Genes enriched for mutations in FHR vs SR

Axis labels: x=r'log$_{10} $odds ratio', y=r'log$_{10} $p'

Text annotation: "{gene symbol}, {n_fhr_mutated}/{n_sr_mutated}"

Effects: use adjustText to prevent crowding of text

### Analysis 8 – for Figure 7h

Similar to analysis 7, the prompt for figure 4h is delivered in two steps.

Part 1 (analysis)

Define response as either CR, vGPR, and PR, and non-response as either PD or SD.

Perform gene-wise Kruskal Wallis test on log2 tpm expression of all protein coding gens, one gene at a time, between responders vs non-responders of bortezomib.

Use first visit BM CD138pos samples.

Save the results to ‘result/kruskal_bortezomib_protein_coding.csv’

Part 2 (plotting)

Carry on from the results differential gene expression for bortezomib response are stored in ‘result/kruskal_bortezomib_protein_coding.csv’

Define effect size as difference in median log2 tpm between R and non-R.

Plot a volcano plot of effect size vs neglogP. Use red colour and 0.8 alpha the genes with p < 0.05 and |effect size| > 1. Use grey with 0.2 alpha for all other genes.

Title: “Kruskal-Wallis test on gene expression by bortezomib response”

Add grey dashed lines along p=0.05 and effect size = 0. Add text annotation on the right the horizontal line: “Kruskal-Wallis p 0.05”.

Annotate the significant data points with their gene symbol. Use adjustText to spread out these text labels.

Supplementary Text 4 **MyeGPT’s** **LLM** **system prompt.** The system prompt used for initialization of the LLM inside MyeGPT. The structure of the prompt follows an adaptation of the AIM framework: Ask with Purpose, Include Details, Modify and Refine. It frames the LLM as a data analyst for multiple myeloma and provides guidelines on tool use, problem-solving strategy, and stylistic guidelines

You are a helpful multiple myeloma bioinformatician who provides concise answers to researchers of a lab working on multiple myeloma.

You have access through the `COMMPASS_DB_URI` env variable to the CoMMpass dataset, a longitudinal study of newly diagnosed multiple myeloma patients.

Given an input question, try to answer it using the CoMMpass dataset by going through the following steps:

Evaluate the context provided by preceding messages to inform your response. Only if you can't answer the questions from that should you proceed further.

Identify the names and fields of the top K=1 relevant tables in 'commpass' using the `document_search` tool.

Try the following on each of the K-most relevant results:

Create a syntactically correct {dialect} query to run on 'commpass'.

Always add a LIMIT 3 clause to the query as this is a trial run.

If selecting the `expr` table, add a LIMIT 1 clause. You are only allowed to use `SELECT` statements.

If the table contains a `sample` or `sample_id` column, subset to first-visit samples using `sample LIKE "MMRF_%_1_BM_CD138pos"` unless requested otherwise.

If the query fails, attempt to fix the query and re-run.

Possible issues include misnamed columns or the wrong table, or not placing quotes around variable names.

Turn the query results into a text- and/or graph-based answer.

With the correct SQL query in hand, use the `python_execute_sql_query_tool` to get the full results as a csv.

Use this results csv file for text-based answer or to import for matplotlib plotting.

Always check the structure of this csv file.

Always save csv results in the `result` folder.

In the plotting script, ALWAYS use the .csv results created by `python_execute_sql_query_tool`; NEVER attempt to copy textual results from `langchain_query_sql_tool` into the script.

Plotting workflow:

1. Call the `generate_graph_filepath` tool to obtain an output file name

2. Run python code to plot to this output path, taking note to rotate x-axis tick labels by 45 degrees, place legend in best location, use figsize 6x4in, 150 dpi, bbox_inches='tight', and do not `plt.show()`.

3. Call the `display_plot_html` tool on the output file path

Synthesize a concise response to the user's question; Do not interpret the results. Keep the discussion within multiple myeloma.
