## Supplementary File 1 for "“MyeGPT: an AI agent for Multiple Myeloma”": supplementary-file-2-question-answer.ipynb

```
# %% [code]
import pandas as pd
pd.options.display.max_columns = 100

# %% [markdown]
## Max visits

# %% [code]
df = pd.read_csv('/Users/jiageng/Projects/myegpt/clindata/MMRF_CoMMpass_IA22_PER_PATIENT_VISIT.tsv',sep='\t')

# %% [code]
df.dropna(subset="VISIT").groupby("PUBLIC_ID").VISIT.nunique()

# %% [code]
df[df.PUBLIC_ID=="MMRF_1140"]

# %% [code]
df.replace(pd.NA, '-').groupby(['BA_LEFT','BA_LEFT2','BA_LEFT3','BA_LEFT4','BA_RIGHT','BA_RIGHT2','BA_RIGHT3','BA_RIGHT4']).size().to_csv('out.csv')

# %% [code]
df.replace(pd.NA, '-').groupby(['BA_LEFT','BA_LEFT2','BA_LEFT3','BA_LEFT4']).size()

# %% [markdown]
## Drug response

# %% [code]
df_trtresp = pd.read_csv('../clindata/MMRF_CoMMpass_IA22_STAND_ALONE_TRTRESP.tsv',sep='\t',encoding='latin')
df_trtresp.head()

# %% [markdown]
### Lenalidomide

# %% [code]
trt_len = df_trtresp['trtname'].str.contains('Lenalidomide')
resist_len = df_trtresp[trt_len]['response'].isin([0])
print('non-responders',len(df_trtresp[trt_len][resist_len].PUBLIC_ID.unique()))
respon_len = df_trtresp[trt_len]['response'].isin([1])
print('responders',len(df_trtresp[trt_len][respon_len].PUBLIC_ID.unique()))

# %% [code]
df_trtresp[trt_len][df_trtresp[trt_len].bestrespsh.isin(['VGPR','CR','PR','sCR'])].groupby('PUBLIC_ID').__len__()

# %% [code]
df_trtresp[trt_len][df_trtresp[trt_len].bestrespsh.isin(['SD','PD'])].groupby('PUBLIC_ID').__len__()

# %% [code]
df_trtresp[trt_len][df_trtresp[trt_len]['bestrespsh'].isna()].groupby('PUBLIC_ID').__len__()

# %% [markdown]
### Pomalidomide

# %% [code]
trt_pom = df_trtresp['trtname'].str.contains('Pomalidomide')

# %% [code]
resist_pom = df_trtresp[trt_pom].response.isin([0])
print('non-responders',len(df_trtresp[trt_pom][resist_pom].PUBLIC_ID.unique()))

respon_pom = df_trtresp[trt_pom].response.isin([1])
print('responders',len(df_trtresp[trt_pom][respon_pom].PUBLIC_ID.unique()))

# %% [code]
df_trtresp[trt_pom][df_trtresp[trt_pom].bestrespsh.isin(['VGPR','CR','PR','sCR'])].groupby('PUBLIC_ID').__len__()

# %% [code]
df_trtresp[trt_pom][df_trtresp[trt_pom].bestrespsh.isin(['SD','PD'])].groupby('PUBLIC_ID').__len__()

# %% [code]
df_trtresp[trt_pom][df_trtresp[trt_pom]['bestrespsh'].isna()].groupby('PUBLIC_ID').__len__()

# %% [markdown]
### Daratumumab

# %% [code]
trt_dara = df_trtresp['trtname'].str.contains('Daratumumab')
print('Option 1: use response column')
respon_data = df_trtresp[trt_dara]['response'].isin([1])
print('responders',len(df_trtresp[trt_dara][respon_data].PUBLIC_ID.unique()))
resist_dara = df_trtresp[trt_dara]['response'].isin([0])
print('non-responders',len(df_trtresp[trt_dara][resist_dara].PUBLIC_ID.unique()))

# %% [code]
respon_dara = df_trtresp[trt_dara][df_trtresp[trt_dara].bestrespsh.isin(['VGPR','CR','PR','sCR'])].groupby('PUBLIC_ID').__len__()
resis_dara = df_trtresp[trt_dara][df_trtresp[trt_dara].bestrespsh.isin(['SD','PD'])].groupby('PUBLIC_ID').__len__()
missing_dara = df_trtresp[trt_dara][df_trtresp[trt_dara]['bestrespsh'].isna()].groupby('PUBLIC_ID').__len__()
print('Option 2: use bestresp/bestrespsh column')
print('responders', respon_dara)
print('non-responders', resis_dara)
print('unknown', missing_dara)

# %% [markdown]
## Induction therapy drug

# %% [code]
df_trtreg = pd.read_csv('/Users/jiageng/Projects/myegpt/clindata/MMRF_CoMMpass_IA22_STAND_ALONE_TREATMENT_REGIMEN.tsv',
            sep='\t',
            encoding='latin')

# %% [code]
df_trtreg.MMTX_TYPE.unique()

# %% [code]
df_trtreg.query('PUBLIC_ID=="MMRF_1564" and MMTX_TYPE!="First line of therapy"')['MMTX_THERAPY'].unique()

# %% [code]
df_trtreg = pd.read_csv('/Users/jiageng/Projects/myegpt/clindata/MMRF_CoMMpass_IA22_STAND_ALONE_TREATMENT_REGIMEN.tsv',
                        sep='\t',
                        encoding='latin')
df_trtreg\
    .query('MMTX_TYPE=="First line of therapy"')\
    .groupby("MMTX_THERAPY")['PUBLIC_ID']\
    .nunique()\
    .sort_values(ascending=False)\
    .head(10)

# %% [markdown]
## Clinical high risk

Relapse within 12 months of induction therapy

# %% [code]
df_trtresp = pd.read_csv('../clindata/MMRF_CoMMpass_IA22_STAND_ALONE_TRTRESP.tsv',sep='\t',encoding='latin')
df_trtresp.query('line==1\
                  and trtstdy==1\
                  and trtgroup==1\
                  and pddy - trtstdy < 365')['PUBLIC_ID'].nunique()

# %% [markdown]
## FHR

* refer to `fhr/01_call_fhr.ipynb`

# %% [markdown]
## Cox regression

# %% [markdown]
### all genes, univariate

# %% [code]
import pandas as pd
import numpy as np
import dotenv
assert dotenv.load_dotenv('.env')
from lifelines import CoxPHFitter
from tqdm import tqdm

# %% [code]
df_cph = pd.read_csv('result/cox_dataset_template_pfs.csv').set_index('PUBLIC_ID')

# %% [code]
df_expr = pd.read_csv('../omicdata/MMRF_CoMMpass_IA22_salmon_geneUnstranded_tpm.csv')

# %% [code]
df_expr1 = df_expr.filter(regex='Gene|MMRF_\\d+_1_BM_CD138pos').set_index('Gene') # first visit samples

df_expr1 = df_expr1.assign(**{col: np.log2(df_expr1[col] + 1) for col in df_expr1.columns})

df_expr1 = df_expr1.apply(lambda x: (x - x.mean()) / x.std(), axis=1)

df_expr1.columns = df_expr1.columns.str.replace('_1_BM_CD138pos','')

# %% [code]
shared_index = sorted(list(set(df_expr1.columns).intersection(set(df_cph.index))))
df_cox = df_cph.loc[shared_index]

# %% [code]

def run_coxph(gene_id):
    ser = df_expr1.loc[gene_id]
    df = df_cox.copy()
    df.loc[shared_index,gene_id] = ser.loc[shared_index]
    return df
    cph = CoxPHFitter()
    cph.fit(df, duration_col='pfscdy', event_col='censpfs')
    df_result = cph.summary
    ser_result = df_result.loc[gene_id]
    ser_result.name = gene_id
    return ser_result

# %% [code]
result = run_coxph('ENSG00000121410')
result

# %% [code]
np.any(pd.isna(result),axis=0)

# %% [code]
result = []

for gene in tqdm(df_expr1.index[:50]):
    try:
        res = run_coxph(gene)
    except Exception as e:
        print(f"Error processing gene {gene}: {e}")
        continue
    result.append(res)


# %% [code]
df_results = pd.DataFrame(result)
df_results.index = [r.name for r in result]
df_results.to_csv('result/cox_ph_os_all_genes.csv')
df_results.shape

# %% [markdown]
### CKS1B

# %% [code]
import pandas as pd
import dotenv
assert dotenv.load_dotenv('.env')

from tools import get_cox_regression_base_data, max_overlapping_segment

# %% [code]
get_cox_regression_base_data(endpoint='os')

# %% [code]
df_cph_os = pd.read_csv('result/cox_dataset_template_os.csv')

# %% [code]
max_overlapping_segment("ENSG00000160710")
cks1b_cn = pd.read_csv('result/gene_level_copy_number_ENSG00000160710.csv',index_col=0)

# %% [code]
df_cph_os = df_cph_os.merge(cks1b_cn[['segment_copy_number_status']], left_on='PUBLIC_ID', right_index=True)

# %% [code]
from lifelines import CoxPHFitter
cph_os =CoxPHFitter()
cph_os.fit(df_cph_os.set_index('PUBLIC_ID'), duration_col='oscdy', event_col='censos')
cph_os.print_summary()

# %% [markdown]
### MYC

# %% [code]
import dotenv
assert dotenv.load_dotenv('.env_eval')
from tools import get_cox_regression_base_data, max_overlapping_segment, convert_gene

# %% [code]
get_cox_regression_base_data(endpoint='pfs')
df_cph_pfs = pd.read_csv('result/cox_dataset_template_pfs.csv')

# %% [code]
max_overlapping_segment(convert_gene('TP53'))

# %% [code]
gene_cn = pd.read_csv('result/gene_level_copy_number_ENSG00000141510.csv',index_col=0)
gene_cn.head()

# %% [code]
df_cph_pfs_gene = df_cph_pfs.merge(gene_cn[['segment_copy_number_status']], left_on='PUBLIC_ID', right_index=True).dropna(subset=['pfscdy'])

# %% [code]
from lifelines import CoxPHFitter
cph_pfs = CoxPHFitter()
cph_pfs.fit(df_cph_pfs_gene.set_index('PUBLIC_ID'), duration_col='pfscdy', event_col='censpfs')
cph_pfs.print_summary()

# %% [markdown]
## CN 1q

# %% [code]
import pandas as pd
import dotenv
assert dotenv.load_dotenv('.env_eval')
from tools import convert_gene, _max_overlapping_segment

# %% [code]
cn_gene_names = ['BCL9', 'ANP32E', 'MCL1', 'RFX5', 'PSMD4', 'ILF2', 'IL6R', 'ADAR', 'CKS1B']
cn_gene_stable_ids = [convert_gene(gene) for gene in cn_gene_names]
cn_gene_dict = dict(zip(cn_gene_names, cn_gene_stable_ids))
cn_gene_dict

# %% [code]
cn_gene_segment_mean = {k : pd.Series(_max_overlapping_segment(v).query('sample.str.contains("_1_BM_CD138pos")')['segment_mean']) for k, v in cn_gene_dict.items()}

# %% [code]
df_cn_gene_segment_mean = pd.concat(cn_gene_segment_mean, axis=1)

# %% [code]
# sanity check to ensure no two columns (genes) have identical CN values
def equal_columns(df):
    cols = df.columns.tolist()
    pairs = []
    for i in range(len(cols)):
        for j in range(i + 1, len(cols)):
            c1, c2 = cols[i], cols[j]
            if df[c1].equals(df[c2]):
                pairs.append((c1, c2))
    return pairs

pairs = equal_columns(df_cn_gene_segment_mean)
print(pairs)

# %% [code]
df_cn_gene_segment_mean

# %% [code]
import numpy as np
corrmat = df_cn_gene_segment_mean.corr()
corrmat.loc[:,::-1]

# %% [code]
np.linalg.det(corrmat)

# %% [markdown]
## TP53 survival

# %% [code]
import pandas as pd
import numpy as np
from lifelines import CoxPHFitter

# %% [code]
dfsurv = pd.read_csv('../clindata/MMRF_CoMMpass_IA22_STAND_ALONE_SURVIVAL.tsv', sep='\t')

# %% [code]
dfmut = pd.read_csv("../omicdata/IGV Downloads-MMRF_CoMMpass_IA22_exome_vcfmerger2_IGV_All_Canonical_NS_Variants.mut",sep='\t')

# %% [code]
px_tp53_mut = dfmut.loc[dfmut.GENE=='TP53']['sample'].str.extract(r'(MMRF_[0-9]+)_')[0].unique()
px_p53_wt = dfsurv.loc[~dfsurv.PUBLIC_ID.isin(px_tp53_mut),'PUBLIC_ID'].unique()

# %% [code]
dfcph = dfsurv[['PUBLIC_ID','oscdy','censos','pfscdy','censpfs']].copy()
dfcph = dfcph.merge(
    pd.DataFrame({'PUBLIC_ID': px_tp53_mut, 'tp53_status': True}),
    on='PUBLIC_ID',
    how='left'
)
dfcph['tp53_status'] = dfcph['tp53_status'].fillna(False)

# %% [code]
dfcphpfs = dfcph.drop(columns=['oscdy','censos']).dropna(subset=['pfscdy'])
dfcphos = dfcph.drop(columns=['pfscdy','censpfs'])

# %% [code]
cphos = CoxPHFitter()
cphos.fit(dfcphos.set_index('PUBLIC_ID'), duration_col='oscdy', event_col='censos')
cphos.print_summary()

# %% [code]
cphpfs = CoxPHFitter()
cphpfs.fit(dfcphpfs.set_index('PUBLIC_ID'), duration_col='pfscdy', event_col='censpfs')
cphpfs.print_summary()

# %% [code]
dfcphpfs_imp = dfcph.copy()
dfcphpfs_imp.loc[dfcph['pfscdy'].isna(),'pfscdy'] = dfcph.loc[dfcph['pfscdy'].isna(),'oscdy']
dfcphpfs_imp.drop(columns=['oscdy','censos'],inplace=True)

# %% [code]
cphpfs_imp = CoxPHFitter()
cphpfs_imp.fit(dfcphpfs_imp.set_index('PUBLIC_ID'), duration_col='pfscdy', event_col='censpfs')
cphpfs_imp.print_summary()

# %% [markdown]
## NSD2 expression

# %% [code]
import pandas as pd
from tools import convert_gene

# %% [code]
dftpm = pd.read_csv("../omicdata/MMRF_CoMMpass_IA22_salmon_geneUnstranded_tpm.tsv",sep='\t')

# %% [code]
gid_nsd2 = convert_gene('NSD2')
gid_nsd2

# %% [code]
tpm_nsd2 = dftpm.loc[dftpm['Gene']==gid_nsd2,:].filter(regex='MMRF_[0-9]+_1_BM_CD138pos').T
tpm_nsd2.columns = ['tpm_nsd2']

# %% [code]
tpm_nsd2.index = tpm_nsd2.index.str.extract(r'(MMRF_[0-9]+)_')[0]

# %% [code]
q4_nsd2 = pd.qcut(tpm_nsd2['tpm_nsd2'], q=4, labels=['Q1','Q2','Q3','Q4'])

# %% [code]
dfsurv = pd.read_csv('../clindata/MMRF_CoMMpass_IA22_STAND_ALONE_SURVIVAL.tsv', sep='\t')
dfos = dfsurv[['PUBLIC_ID','oscdy','censos']]
dfpfs = dfsurv[['PUBLIC_ID','pfscdy','censpfs']].dropna(subset=['pfscdy'])

# %% [code]
dfos_nsd2 = dfos.merge(q4_nsd2, left_on='PUBLIC_ID', right_index=True)
dfpfs_nsd2 = dfpfs.merge(q4_nsd2, left_on='PUBLIC_ID', right_index=True)

# %% [code]
from lifelines import KaplanMeierFitter

# %% [markdown]
Median OS by NSD2 expression quantile

# %% [code]
for label in ['Q1','Q2','Q3','Q4']:
    kmf = KaplanMeierFitter()
    mask = dfos_nsd2['tpm_nsd2']==label
    kmf.fit(dfos_nsd2.loc[mask,'oscdy'], event_observed=dfos_nsd2.loc[mask,'censos'], label=label)
    
    print(label, kmf.median_survival_time_)

# %% [markdown]
Median PFS by NSD2 expression quantile

# %% [code]
for label in ['Q1','Q2','Q3','Q4']:
    kmf = KaplanMeierFitter()
    mask = dfpfs_nsd2['tpm_nsd2']==label
    kmf.fit(dfpfs_nsd2.loc[mask,'pfscdy'], event_observed=dfpfs_nsd2.loc[mask,'censpfs'], label=label)
    
    print(label, kmf.median_survival_time_)
```
