## Supplementary File 2 for "“MyeGPT: an AI agent for Multiple Myeloma”": supplementary-file-4-call-fhr.ipynb

```
# %% [code]
import pandas as pd
pd.set_option('display.max_columns', 50)
pd.set_option('display.max_rows', 50)

# %% [code]
import os
os.chdir('/Users/jiageng/Projects/fhr/pipeline')
os.getcwd()

# %% [code]
file_trtresp = '../data/CoMMpass_IA22_FlatFiles/MMRF_CoMMpass_IA22_STAND_ALONE_TRTRESP.tsv'
df_trtresp = pd.read_csv(file_trtresp, sep='\t', encoding='cp1252')
file_clin = '../data/CoMMpass_IA22_FlatFiles/MMRF_CoMMpass_IA22_PER_PATIENT.tsv'
df_clin = pd.read_csv(file_clin, sep='\t')

# %% [code]
names_all = df_clin.PUBLIC_ID.unique().tolist()
print(len(names_all))

# %% [markdown]
Identification of refractory to induction therapy or early relapse

# %% [code]
df_trtresp.sample(n=3)

# %% [markdown]
Changes on 2 Mar 2025

Tighten threshold for time between treatment initiation and progression to 12 months from 18 months

Number of FHR drops from 186 to 154

# %% [code]
# Clinical high risk
# 1. Number of patients with non-responsive to induction therapy (line 1, treatment group 1)
# 2. Or with progression within 12 months of start of any treatment, 
# 3. Or with progression within 12 months of autologous stem cell transplant
names_clinhr = \
df_trtresp.query("\
    (bestresp in ['Stable Disease','Progressive Disease'] and line==1 and trtgroup==1) or \
    (line==1 and trtgroup==1 and pddy > trtstdy and pddy - trtstdy < 365) or \
    (bmtx_rec==1 and  bmtx_type=='Stem cell, Autologous' and pddy > bmtx_day and pddy - bmtx_day < 365)")\
        .PUBLIC_ID.unique().tolist()
len(names_clinhr)

# %% [code]
# Clinical high risk
# 1. Number of patients with non-responsive to induction therapy (line 1, treatment group 1)
non_responders = \
    df_trtresp.query("\
    (bestresp in ['Stable Disease','Progressive Disease'] and line==1 and trtgroup==1)\
    ").PUBLIC_ID.unique().tolist()
non_responders.__len__()

# %% [code]
# 2. Number of refractory patients
refractory = \
    df_trtresp.query("\
    (line==1 and trtgroup==1 and pddy > trtstdy and pddy - trtstdy < 365) or \
    (bmtx_rec==1 and  bmtx_type=='Stem cell, Autologous' and pddy > bmtx_day and pddy - bmtx_day < 365)")\
        .PUBLIC_ID.unique().tolist()
refractory.__len__()

# %% [code]
import shutil
# Build clinical high-risk flags and write to CSV
public_ids = sorted(set(non_responders).union(refractory))
df_clin_hr = pd.DataFrame({
    "PUBLIC_ID": public_ids,
    "non_responder": [1 if pid in non_responders else 0 for pid in public_ids],
    "refractory": [1 if pid in refractory else 0 for pid in public_ids],
})
df_clin_hr.to_csv("clinical_high_risk.csv", index=False)
shutil.copy("clinical_high_risk.csv", "/Users/jiageng/Projects/myegpt/src/result/human/clinical_high_risk.csv")

# %% [code]
tmp = pd.read_csv('/Users/jiageng/Projects/fhr/data/SeqFISH Files_MMRF_CoMMpass_IA16a_LongInsert_Canonical_Ig_Translocations.txt', sep='\t')\
    .query("Study_Visit_iD.str.endswith('_BM')")\
    .assign(PUBLIC_ID=lambda x: x.Study_Visit_iD.apply(lambda x: '_'.join(x.split('_')[:2])))\
    .sort_values('Study_Visit_iD')\
    .groupby("PUBLIC_ID")\
    .head(n=1)
print(tmp.PUBLIC_ID.unique().size)
tmp.groupby(tmp['Study_Visit_iD'].str.split('_').str[2]).size()

# %% [code]
tmp = pd.read_csv('/Users/jiageng/Projects/fhr/data/SeqFISH Files_MMRF_CoMMpass_IA16a_RNAseq_Canonical_Ig_Translocations.txt', sep='\t')\
    .query("Specimen_ID.str.endswith('_BM')")\
    .assign(PUBLIC_ID=lambda x: x.Specimen_ID.apply(lambda x: '_'.join(x.split('_')[:2])))\
    .sort_values('Specimen_ID')\
    .groupby("PUBLIC_ID")\
    .head(n=1)

print(tmp.PUBLIC_ID.unique().size)
tmp.groupby(tmp['Specimen_ID'].str.split('_').str[2]).size()

# %% [markdown]
Changes on 6 Nov 2025: remove the requirement for samples to come from bone marrow plasma cell 

This adds two Peripheral Blood samples to the individuals with high-risk translocations

The numbers are now 906 | 116 | 37 | 152


# %% [code]
# translocation-cyclin D classifications

use_RNAseq = False

if use_RNAseq:
    file_tc = '../data/SeqFISH Files_MMRF_CoMMpass_IA16a_RNAseq_Canonical_Ig_Translocations.txt'
    df_tc = pd.read_csv(file_tc, sep='\t')\
        .query("Specimen_ID.str.endswith('_BM')")\
        .assign(PUBLIC_ID=lambda x: x.Specimen_ID.apply(lambda x: '_'.join(x.split('_')[:2])))\
        .sort_values('Specimen_ID')\
        .groupby("PUBLIC_ID")\
        .head(n=1)\
        .filter(regex='PUBLIC_ID|_Call')  # RNAseq 
    names_tc = df_tc.query('RNASeq_FGFR3_Call == 1 or RNASeq_MAF_Call == 1')['PUBLIC_ID'].unique() 
else:
    # use long insert WGS
    file_tc = '../data/SeqFISH Files_MMRF_CoMMpass_IA16a_LongInsert_Canonical_Ig_Translocations.txt'
        # .query("Study_Visit_iD.str.endswith('_BM')")\
    df_tc = pd.read_csv(file_tc, sep='\t')\
        .assign(PUBLIC_ID=lambda x: x.Study_Visit_iD.apply(lambda x: '_'.join(x.split('_')[:2])))\
        .sort_values('Study_Visit_iD')\
        .groupby("PUBLIC_ID")\
        .head(n=1)\
        .filter(regex='PUBLIC_ID|_CALL') 
    names_t4_14 = df_tc.query('SeqWGS_WHSC1_CALL == 1')['PUBLIC_ID'].unique()
    names_t14_16 = df_tc.query('SeqWGS_MAF_CALL == 1')['PUBLIC_ID'].unique()
    names_tc = df_tc.query('SeqWGS_WHSC1_CALL == 1 or SeqWGS_MAF_CALL == 1')['PUBLIC_ID'].unique() 

print('Patients with WGS canonical Ig info',len(df_tc.PUBLIC_ID.unique()))

# t(4;14) FGFR3 overexpression or t(14;16) c-MAF overexpression
print('Patients with t(4;14) or t(14;16)',names_tc.__len__())
print('Patients with t(4;14)',names_t4_14.__len__())
print('Patients with t(14;16)',names_t14_16.__len__())

# %% [code]
ser_hr_transloc = sorted(set(names_t4_14).union(set(names_t14_16)))
with open("hr_translocations.csv", "w") as f:
    f.write("PUBLIC_ID\n")
    for pid in ser_hr_transloc:
        f.write(f"{pid}\n")
shutil.copy("hr_translocations.csv", "/Users/jiageng/Projects/myegpt/src/result/human/hr_translocations.csv")

# %% [markdown]
## TP53 double hit

Changes on 2 Mar 2025

Increase segment mean threshold for TP53 deletion from -0.2 to -0.5

Increase segment mean threshold for 1q gain from 0.2 to 0.38

Number with biallelic TP53 drops to 36 from 39

Changes on 11 Jan 2026

remove requirement to come from first visit samples

# %% [code]
# bi-allelic TP53 deletion
# exome CN only has 1051 samples, while genome CN has 1121 samples
# 26 Jul - subset to first visit samples
df_fish = '../data/SeqFISH Files_MMRF_CoMMpass_IA22_genome_gatk_cna_seqFISH.tsv'
df_fish = pd.read_csv(df_fish, sep='\t').query(r"SAMPLE.str.contains('^MMRF_\\d+_1_BM_CD138pos$', regex=True)").assign(PUBLIC_ID=lambda x: x['SAMPLE'].str.extract(r'(MMRF_\d{4})'))
print('df_fish',df_fish.shape)

# TP53 deletion
names_del_tp53 = df_fish.loc[-0.5 > df_fish['SeqWGS_Cp_TP53'],'PUBLIC_ID'].unique() # 65

# Double copy TP53 deletion
# There are no individuals with -2 > log2FC 
names_deep_del_tp53 = df_fish.loc[-1.5 > df_fish['SeqWGS_Cp_TP53'],'PUBLIC_ID'].unique() # 0
assert len(names_deep_del_tp53) == 0

# TP53 mutations
file_mut = '../data/IGV Downloads_MMRF_CoMMpass_IA22_exome_vcfmerger2_IGV_All_Canonical_NS_Variants.mut'
df_mut = pd.read_csv(file_mut, sep='\t')
df_mut = df_mut[df_mut['sample'].str.contains('MMRF_\\d+_\\d+_BM_CD138pos')].assign(PUBLIC_ID=lambda x: x['sample'].str.extract(r'(MMRF_\d{4})'))
# df_mut = df_mut[df_mut['sample'].str.contains('MMRF_\\d+_1_BM_CD138pos')].assign(PUBLIC_ID=lambda x: x['sample'].str.extract(r'(MMRF_\d{4})'))

names_mut_tp53 = df_mut.query('chr=="chr17" and GENE=="TP53"')['PUBLIC_ID'].unique()

# TP53 LOH
# 26 Jul - subset to first visit samples
file_baf = '../data/Loss of Heterozygosity Files_MMRF_CoMMpass_IA22_exome_gatk_baf.seg'
df_baf = pd.read_csv(file_baf, sep='\t').query(r"SAMPLE.str.contains('^MMRF_\\d+_\\d+_BM_CD138pos$', regex=True)").assign(PUBLIC_ID=lambda x: x['SAMPLE'].str.extract(r'(MMRF_\d{4})'))
# df_baf = pd.read_csv(file_baf, sep='\t').query(r"SAMPLE.str.contains('^MMRF_\\d+_1_BM_CD138pos$', regex=True)").assign(PUBLIC_ID=lambda x: x['SAMPLE'].str.extract(r'(MMRF_\d{4})'))
TP53_start=7661779
TP53_end=7687564
# LEN=(TP53_end-TP53_start+1)/2
LEN=1000
# select where (Start, End) overlaps with (TP53_start, TP53_end) by at least @LEN positions
df_baf_tp53 = df_baf.query("Chromosome=='chr17' and ((Start < @TP53_start and End + @LEN > @TP53_start) or (Start < @TP53_end - @LEN and End >= @TP53_end) or (Start > @TP53_start and End < @TP53_end and (End - Start) >= @LEN))")
# LOH defined as B-allele frequency of <0.25 (since it ranges from 0 to 0.5)
names_loh_tp53 = df_baf_tp53.loc[df_baf_tp53['Segment_Mean'] < 0.25, 'PUBLIC_ID'].unique() # 104

# Double Hit TP53 events
# deciding not to include deletion + LOH as a bi-allelic event
biallelic_tp53_type1 = set(names_mut_tp53).intersection(set(names_del_tp53))
biallelic_tp53_type2 = set(names_mut_tp53).intersection(set(names_loh_tp53))
# biallelic_tp53_type3 = set(names_del_tp53).intersection(set(names_loh_tp53))
biallelic_tp53 = biallelic_tp53_type1.union(biallelic_tp53_type2) #.union(biallelic_tp53_type3)

print('Patients with TP53 BAF info',df_baf_tp53['PUBLIC_ID'].unique().__len__()) # 974
print('Patients with TP53 copy number info',df_fish['PUBLIC_ID'].unique().__len__()) # 892
print('Patients with TP53 mutation info',df_mut['PUBLIC_ID'].unique().__len__()) # 974
print(f"Patients with known bi-allelic loss of TP53: {len(biallelic_tp53)} ({len(biallelic_tp53)/892*100:.2f}%)")
print(f"Breakdown: type 1: {len(biallelic_tp53_type1)}, type 2: {len(biallelic_tp53_type2)}")


# %% [code]
print(len(names_mut_tp53))
print(len(names_del_tp53))
print(len(names_loh_tp53))
print(len(set(biallelic_tp53_type1).union(set(biallelic_tp53_type2))))

# %% [code]
doublehit_pid = sorted(set(biallelic_tp53_type1).union(set(biallelic_tp53_type2)))
doublehit_df = pd.DataFrame({
    "PUBLIC_ID": doublehit_pid,
    "mutation_and_deletion": [1 if pid in biallelic_tp53_type1 else 0 for pid in doublehit_pid],
    "mutation_and_loh": [1 if pid in biallelic_tp53_type2 else 0 for pid in doublehit_pid],
})
doublehit_df.to_csv("biallelic_tp53_loss.csv", index=False)
shutil.copy("biallelic_tp53_loss.csv", "/Users/jiageng/Projects/myegpt/src/result/human/biallelic_tp53_loss.csv")

# %% [markdown]
## 1q Gain and ISS stage 3
Increase Gain1q segment mean criteria from 0.2 to 0.38.

Number of Gain1q drops from 385 to 309.

Number of ISS III and Gain1q drops from 110 to 98.

# %% [code]
# 1q gain and ISS3
names_iss3 = df_clin.loc[df_clin['D_PT_iss']==3,'PUBLIC_ID'].unique() # 311
names_gain1q = df_fish.loc[(df_fish['SeqWGS_Cp_1q21'] > 0.38) & (df_fish['SAMPLE'].str.contains('MMRF_\\d+_1_BM_CD138pos')),'PUBLIC_ID'].unique() # 385 # SeqExome
names_gain1q_iss3 = set(names_iss3).intersection(set(names_gain1q)) # 110
print(len(names_iss3))
print(len(names_gain1q))
print(len(names_gain1q_iss3))

# %% [code]
with open("gain1q_iss3.csv", "w") as f:
    f.write("PUBLIC_ID\n")
    for pid in sorted(names_gain1q_iss3):
        f.write(f"{pid}\n")
shutil.copy("gain1q_iss3.csv", "/Users/jiageng/Projects/myegpt/src/result/human/iss3_gain1q.csv")

# %% [code]
# rescue the missing TP53 deletion info using BAF info

df_baf_fish = df_baf_tp53.merge(df_fish.drop(columns=['SAMPLE']),how='outer',on='PUBLIC_ID')
df_baf_only = df_baf_fish[df_baf_fish['SeqWGS_Cp_17p13'].isna()]

# these have two copies of TP53
names_tp53_baf_derived_normal_cn = set(df_baf_only[df_baf_only['Segment_Mean'] > 0.45].PUBLIC_ID)

print(len(names_tp53_baf_derived_normal_cn))

# %% [code]
df_baf_fish.shape

# %% [code]
df_baf_only.shape

# %% [markdown]
## FHR calling simple method

This is not exactly accurate because individuals with missing data are considered lacking the features

e.g. Those without TP53 copy number data (FISH) are assumed to be TP53-copy number neutral

# %% [markdown]
Update 6 Nov 2025: use this as the "correct" genomic high risk numbers

# %% [markdown]
11 Jan 2026 - use this as human answer. Do not adjust for BAF recoverable TP53 cases

# %% [code]
# FHR calling pipeline
names_ghr = set(names_tc).union(set(names_gain1q_iss3)).union(set(biallelic_tp53))
print('GHR',len(names_ghr))
names_fhr = set(names_clinhr).difference(names_ghr)
print('FHR',len(names_fhr))
names_sr = set(names_all).difference(names_ghr).difference(names_fhr)
print('SR',len(names_sr))

# %% [code]
genomic_hr = pd.DataFrame({
    'PUBLIC_ID': sorted(names_ghr),
    'gain1q_iss3': [1 if pid in names_gain1q_iss3 else 0 for pid in sorted(names_ghr)],
    'biallelic_tp53_loss': [1 if pid in biallelic_tp53 else 0 for pid in sorted(names_ghr)],
    'hr_translocation': [1 if pid in names_tc else 0 for pid in sorted(names_ghr)],
})
genomic_hr.to_csv('genomic_hr.csv', index=False)
shutil.copy("genomic_hr.csv", "/Users/jiageng/Projects/myegpt/src/result/human/genomic_hr.csv")

# %% [code]
with open("functional_hr.csv","w") as f:
    f.write("PUBLIC_ID\n")
    for pid in sorted(names_fhr):
        f.write(f"{pid}\n")
shutil.copy("functional_hr.csv", "/Users/jiageng/Projects/myegpt/src/result/human/functional_hr.csv")

# %% [code]
with open("standard_risk.csv","w") as f:
    f.write("PUBLIC_ID\n")
    for pid in sorted(names_sr):
        f.write(f"{pid}\n")
shutil.copy("standard_risk.csv", "/Users/jiageng/Projects/myegpt/src/result/human/standard_risk.csv")

# %% [markdown]
# Data frame of constituent FHR features

# %% [markdown]
FHR features in OncoPrint format
public_id feature_1_yes/no/missing ... feature_n_yes/no/missing (1=yes, 0=no, -1=missing)

# %% [code]
names_refractory = df_trtresp.query("(bestrespsh in ['SD','PD'] and line==1 and trtgroup==1)").PUBLIC_ID.unique()
names_relapse = df_trtresp.query("(line==1 and trtgroup==1 and pddy > trtstdy and pddy - trtstdy < 365) or (bmtx_rec==1 and bmtx_type=='Stem cell, Autologous' and pddy > bmtx_day and pddy - bmtx_day < 365)").PUBLIC_ID.unique()
names_clinhr = set(names_refractory).union(set(names_relapse)).intersection(set(names_all))

names_tc_all = df_tc.PUBLIC_ID.unique()
names_cn_all = set(df_fish.PUBLIC_ID).union(set(names_tp53_baf_derived_normal_cn))
names_baf_all = df_baf.PUBLIC_ID.unique()
names_mut_all = df_mut.PUBLIC_ID.unique()

names_iss_all = df_clin.dropna(subset='D_PT_iss').PUBLIC_ID.unique()

names_tp53_all = set(names_cn_all).union(set(names_mut_all)).union(set(names_baf_all))
len(names_tp53_all)
names_tp53_mut_del = set(names_mut_tp53).intersection(set(names_del_tp53))
names_tp53_mut_loh = set(names_mut_tp53).intersection(set(names_loh_tp53))
names_tp53_biallelic = set(names_tp53_mut_del).union(set(names_tp53_mut_loh))
data = {
    'Refractory': [1 if name in names_refractory else 0 for name in names_all],
    'Early relapse': [1 if name in names_relapse else 0 for name in names_all],
    't(4;14)/t(14;16)': [1 if name in names_tc else 0 if name in names_tc_all else -1 for name in names_all],
    'Double Hit TP53': [1 if name in names_tp53_biallelic else 0 if name in names_tp53_all else -1 for name in names_all],
    'TP53 NS Mut': [1 if name in names_mut_tp53 else 0 if name in names_mut_all else -1 for name in names_all],
    'TP53 Del': [1 if name in names_del_tp53 else 0 if name in names_cn_all else -1 for name in names_all],
    'TP53 LoH': [1 if name in names_loh_tp53 else 0 if name in names_baf_all else -1 for name in names_all],
    'ISS III': [1 if name in names_iss3 else 0 if name in names_iss_all else -1 for name in names_all],
    'Gain1q': [1 if name in names_gain1q else 0 if name in names_cn_all else -1 for name in names_all]
}

df_fhr = pd.DataFrame(data,index=names_all)
df_fhr.index.name='PUBLIC_ID'

# %% [code]
df_fhr

# %% [code]
# individuals with data present across all genomic features
names_ghr_all = set(names_tc_all).intersection(set(names_cn_all)).intersection(set(names_baf_all)).intersection(set(names_mut_all)).intersection(set(names_iss_all).union(names_gain1q))
names_ghr = set(names_tc).union(set(names_tp53_biallelic)).union((set(names_iss3).union(names_gain1q))).intersection(names_ghr_all)

names_fhr = set(names_clinhr).difference(names_ghr)

# %% [code]
# get a quick count of how many FHR patients are in the dataset
labels_fhr = [1 if name in names_fhr else -1 for name in names_all]
pd.Series(labels_fhr).value_counts()

# %% [code]
labels_all = [2 if name in names_fhr else 1 if (name in names_ghr and name in names_ghr_all) else 0 for name in names_all]
pd.Series(labels_all).value_counts()

# %% [code]
from matplotlib import pyplot as plt
import numpy as np
from matplotlib.colors import ListedColormap

# for oncoplot side axes
from mpl_toolkits.axes_grid1.axes_divider import make_axes_locatable
from matplotlib.patches import Patch


# %% [code]
# Order the samples using the oncoplot algorithm
def oncoplot_ordering(data,extra_covariates=[],prioritize_covariates=False):
    # Define a recursive function to order the data
    def recursive_ordering(data, depth=0):
        if depth >= data.shape[1]:
            return data.index.tolist()
        
        # Order columns by column-wise sum
        primary_col = ordered_columns[depth]
                
        # The remaining are IGH partner columns
        # Divide observations into groups based on the primary column
        
        ordered_indices = []
        
        for value in sorted(data[primary_col].unique())[::-1]:
            # subset to observations with the same value
            group_v = data[data[primary_col] == value]
            # Recursively order within each group
            ordered_indices_v = recursive_ordering(group_v, depth + 1)
            # Combine the ordered indices
            ordered_indices.extend(ordered_indices_v)
        
        return ordered_indices
    
    # Start the recursive ordering
    # Order by number of carriers
    # ordered_columns = (data==1).sum(axis=0).sort_values(ascending=False).index
    heatmap_data = data.drop(columns=extra_covariates)
    ordered_heatmap_columns = (heatmap_data==1).sum(axis=0).sort_values(ascending=False).index
    
    # Order additional columns appear last
    if prioritize_covariates:    
        ordered_columns = pd.Index(extra_covariates).append(ordered_heatmap_columns)
    else:
        ordered_columns = ordered_heatmap_columns.append(pd.Index(extra_covariates))
    
    ordered_indices = recursive_ordering(data.loc[:, ordered_columns])
    return data.loc[ordered_indices, ordered_columns]


# %% [code]
df_fhr_ordered = oncoplot_ordering(df_fhr)

# %% [code]
df_fhr_ordered.shape

# %% [code]
(df_fhr_ordered==1).sum()

# %% [code]
import seaborn as sns
fig,ax=plt.subplots()
fig.set_size_inches(16,8)
ax=sns.heatmap(df_fhr_ordered.T, cmap=ListedColormap(['#AFAFAF','#FAFAFA','red']), cbar=False, xticklabels=False,ax=ax)
ax.set_title('FHR contributing features')

# add black frame around heatmap
for spine in ax.spines.values():
    spine.set_visible(True)
    
# Create legend for main heatmap
main_legend_elements = [
    Patch(facecolor='white', edgecolor='black', label='Non-carrier'),
    Patch(facecolor='red', edgecolor='black', label='Carrier'),
    Patch(facecolor='grey', edgecolor='black', label='Missing'),
]
ax.legend(handles=main_legend_elements, title='Others', bbox_to_anchor=(1, 0.5), loc='upper left', frameon=True)

fig.savefig('../fhr-features-only-heatmap-full-1May26.svg')

# %% [markdown]
# Correct case by case FHR calling method

# %% [markdown]
New method which does not use set to assign FHR, considers case by case
* 1 May: relax criterion. Unknown = considered absent for the feature
* This will give 114, 248, and 781 cases
* In MyeGPT, GHR has 250 cases because of 2 PB (peripheral blood) samples. It is 248 here because we only consider Bone Marrow samples

# %% [code]
df_fhr_ann = df_fhr.copy()
df_fhr_ann.loc[:,'risk'] = pd.Series(dtype=int)

# Manually call FHR, GHR, and SR
for i,x in df_fhr_ann.iterrows():
    # i is MMRF_1014
    if (x['Refractory']==1 or x['Early relapse']==1) \
            and x['t(4;14)/t(14;16)']<=0 \
            and x['Double Hit TP53']<=0 \
            and (x['ISS III']<=0 or x['Gain1q']<=0):
                df_fhr_ann.loc[i,'risk'] = 2 # 'FHR'
    elif (x['Refractory']==0 and x['Early relapse']==0) \
            and x['t(4;14)/t(14;16)']<=0 \
            and x['Double Hit TP53']<=0 \
            and (x['ISS III']<=0 or x['Gain1q']<=0):
                df_fhr_ann.loc[i,'risk'] = 0 # 'SR'
    elif (x['t(4;14)/t(14;16)']>0 or x['Double Hit TP53']>0 or (x['ISS III']>0 and x['Gain1q']>0)):
        df_fhr_ann.loc[i,'risk'] = 1 # 'GHR'
    else:
        df_fhr_ann.loc[i,'risk'] = -1 # 'NA'

df_fhr_ann['risk'] = df_fhr_ann['risk'].astype(int)
df_fhr_ann['risk'].value_counts()

# %% [code]
# Create ordered data with secondary labels
df_fhr_ann_ordered = oncoplot_ordering(df_fhr_ann, extra_covariates=['risk'],prioritize_covariates=False)

ordered_data = df_fhr_ann_ordered.drop(columns='risk')
ordered_colnames = ordered_data.columns
# Create OncoPrint
plt.clf(); 
fig,ax = plt.subplots()
fig.set_size_inches(16,8)

xlim = ax.set_xlim(0,ordered_data.shape[0])
ylim = ax.set_ylim(0,ordered_data.shape[1])

# instead of using axes.imshow, use axes.add_patch to draw rectangles
for i in range(ordered_data.shape[0]):
    for j in range(ordered_data.shape[1]):
        # draw a thin rectangle for each 1 in the data
        if ordered_data.iloc[i, j] == 1:
            ax.add_patch(
                plt.Rectangle(
                    (i, ordered_data.shape[1]-1-j), 1, 1, linewidth=0, color='#FF2222'
                )
            )
        elif ordered_data.iloc[i,j] == -1:
            ax.add_patch(
                plt.Rectangle(
                    (i, ordered_data.shape[1]-1-j), 1, 1, linewidth=0, color='#CCCCCC'
                )
            )

# offset y-axis ticks to center them
ax.set_yticks(np.arange(len(ordered_colnames))+0.5)
# need to reverse the order of the labels this time
ax.set_yticklabels(ordered_colnames[::-1], rotation=0)

# Create legend for main heatmap
main_legend_elements = [
    Patch(facecolor='white', edgecolor='black', label='Non-carrier'),
    Patch(facecolor='#FF2222', edgecolor='black', label='Carrier'),
    Patch(facecolor='#CCCCCC', edgecolor='black', label='Missing'),
]
ax.legend(handles=main_legend_elements, title='Matrix', bbox_to_anchor=(1, 0.5), loc='upper left', frameon=True)

# Add secondary annotations
ax = make_axes_locatable(ax)

# Create a new axes for the secondary annotation
ax_fhr = ax.append_axes("top", size="7%", pad="2%")

# Define a colormap for the risk types
# fhr_cmap = ListedColormap(['#AAAAAA','white','#FFAAAA','#FF2222'])
# 1 May 26: if there is no 'Missing', then remove the grey color
fhr_cmap = ListedColormap(['white','#FFAAAA','#FF2222'])


# Plot the 1D heatmap for risk types
fhr_values = df_fhr_ann_ordered['risk'].values

ax_fhr.imshow(fhr_values[np.newaxis,:], aspect='auto', cmap=fhr_cmap)

# Remove ticks
ax_fhr.set_xticks([])
ax_fhr.set_yticks([])


# Add y-axis label for secondary annotation
ax_fhr.set_ylabel('Risk', rotation=0, labelpad=5, va='center', ha='right')

# Create legend for FHR
fhr_legend_elements = [
    Patch(facecolor='white', edgecolor='black', label='SR'),
    Patch(facecolor='#FFAAAA', edgecolor='black', label='GHR'),
    Patch(facecolor='#FF2222', edgecolor='black', label='FHR'),
    Patch(facecolor='#AAAAAA', edgecolor='black', label='Missing'),
]
ax_fhr.legend(handles=fhr_legend_elements, title='Risk', bbox_to_anchor=(1, -3), loc='upper left', frameon=True)

plt.title('Constituent features for risk classification')

fig.savefig('../fhr-heatmap-full-ann.1May26.svg')

# %% [markdown]
Remove early relapse and refractory from the main heatmap and show them as covariates

# %% [code]
# Create ordered data with covariates
covariates = ['risk','Early relapse','Refractory']
df_fhr_ann_ordered = oncoplot_ordering(df_fhr_ann, extra_covariates=covariates,prioritize_covariates=False)

ordered_data = df_fhr_ann_ordered.drop(columns=covariates)
ordered_colnames = ordered_data.columns

# Create OncoPrint with side axes
plt.clf(); 
fig,ax = plt.subplots()
fig.set_size_inches(16,8)

xlim = ax.set_xlim(0,ordered_data.shape[0])
ylim = ax.set_ylim(0,ordered_data.shape[1])

color_red = "#FF2222"
color_grey = "#CCCCCC"
color_pink = "#FFAAAA"

# instead of using axes.imshow, use axes.add_patch to draw rectangles
for i in range(ordered_data.shape[0]):
    for j in range(ordered_data.shape[1]):
        # draw a thin rectangle for each 1 in the data
        if ordered_data.iloc[i, j] == 1:
            ax.add_patch(
                plt.Rectangle(
                    (i, ordered_data.shape[1]-1-j), 1, 1, linewidth=0, color=color_red
                )
            )
        elif ordered_data.iloc[i,j] == -1:
            ax.add_patch(
                plt.Rectangle(
                    (i, ordered_data.shape[1]-1-j), 1, 1, linewidth=0, color=color_grey
                )
            )

# offset y-axis ticks to center them
ax.set_yticks(np.arange(len(ordered_colnames))+0.5)
# need to reverse the order of the labels this time
ax.set_yticklabels(ordered_colnames[::-1], rotation=0)


# Create legend for main heatmap
main_legend_elements = [
    Patch(facecolor='white', edgecolor='black', label='No'),
    Patch(facecolor=color_red, edgecolor='black', label='Yes'),
    Patch(facecolor=color_grey, edgecolor='black', label='Missing'),
]
ax.legend(handles=main_legend_elements, title='Matrix', bbox_to_anchor=(1, 0.5), loc='upper left', frameon=False)

# Add secondary annotations
ax = make_axes_locatable(ax)

# Create a new axes for the secondary annotation
ax_fhr = ax.append_axes("top", size="7%", pad="2%")
ax_early = ax.append_axes("top", size="7%", pad="2%") # early relapse
ax_refrac = ax.append_axes("top", size="7%", pad="2%") # refractory

# Define a colormap for the risk types
cmap_fhr = ListedColormap([color_grey,'white',color_pink,color_red])
cmap_early = ListedColormap(['white',color_red])
cmap_refrac = ListedColormap(['white',color_red])

# Plot the 1D heatmap for risk types
values_fhr = df_fhr_ann_ordered['risk'].values
values_early = df_fhr_ann_ordered['Early relapse'].values
values_refrac = df_fhr_ann_ordered['Refractory'].values

ax_fhr.imshow(values_fhr[np.newaxis,:], aspect='auto', cmap=cmap_fhr)
ax_early.imshow(values_early[np.newaxis,:], aspect='auto', cmap=cmap_early)
ax_refrac.imshow(values_refrac[np.newaxis,:], aspect='auto', cmap=cmap_refrac)

# Remove ticks
ax_fhr.set_xticks([])
ax_fhr.set_yticks([])
ax_early.set_xticks([])
ax_early.set_yticks([])
ax_refrac.set_xticks([])
ax_refrac.set_yticks([])

# Add y-axis label for secondary annotation
ax_fhr.set_ylabel('Risk', rotation=0, labelpad=5, va='center', ha='right')
ax_early.set_ylabel('Early relapse', rotation=0, labelpad=5, va='center', ha='right')
ax_refrac.set_ylabel('Refractory', rotation=0, labelpad=5, va='center', ha='right')

# Create legend for FHR
legend_elements_fhr = [
    Patch(facecolor='white', edgecolor='black', label='SR'),
    Patch(facecolor=color_pink, edgecolor='black', label='GHR'),
    Patch(facecolor=color_red, edgecolor='black', label='FHR'),
    Patch(facecolor=color_grey, edgecolor='black', label='Missing'),
]
ax_fhr.legend(handles=legend_elements_fhr, title='Risk', bbox_to_anchor=(1, -2), loc='upper left', frameon=False)

# Create legend for Early relapse
legend_elements_early = [
    Patch(facecolor='white', edgecolor='black', label='No'),
    Patch(facecolor=color_red, edgecolor='black', label='Yes'),
]
ax_early.legend(handles=legend_elements_early, title='Early relapse', bbox_to_anchor=(1, -1), loc='upper left', frameon=False)

# Create legend for Refractory
legend_elements_refrac = [
    Patch(facecolor='white', edgecolor='black', label='No'),
    Patch(facecolor=color_red, edgecolor='black', label='Yes'),
]
ax_refrac.legend(handles=legend_elements_refrac, title='Refractory', bbox_to_anchor=(1, 0), loc='upper left', frameon=False)

plt.title('Constituent features for risk classification')

fig.savefig('../fhr-heatmap-full-ann-rr.1May26.svg')

# %% [markdown]
Order by the heatmap by the risk classification first
Looks weird because the columns are ordered differently

# %% [code]
# Create ordered data, ordered by risk classification first, then by oncoplot ordering within each risk group
# Group by risk category and apply oncoplot ordering within each group
risk_groups = df_fhr_ann.groupby('risk')
ordered_indices = []

for risk_val in sorted(df_fhr_ann['risk'].unique())[::-1]:
    group = df_fhr_ann[df_fhr_ann['risk'] == risk_val]
    # Apply oncoplot ordering within each risk group
    group_ordered = oncoplot_ordering(group.drop(columns='risk'), extra_covariates=[], prioritize_covariates=False)
    ordered_indices.extend(group_ordered.index.tolist())

df_fhr_ann_ordered = df_fhr_ann.loc[ordered_indices]

ordered_data = df_fhr_ann_ordered.drop(columns=['risk'])
ordered_colnames = ordered_data.columns

# Create OncoPrint
plt.clf(); 
fig,ax = plt.subplots()
fig.set_size_inches(16,8)

xlim = ax.set_xlim(0,ordered_data.shape[0])
ylim = ax.set_ylim(0,ordered_data.shape[1])

color_red = "#FF2222"
color_grey = "#CCCCCC"
color_pink = "#FFAAAA"

# instead of using axes.imshow, use axes.add_patch to draw rectangles
for i in range(ordered_data.shape[0]):
    for j in range(ordered_data.shape[1]):
        # draw a thin rectangle for each 1 in the data
        if ordered_data.iloc[i, j] == 1:
            ax.add_patch(
                plt.Rectangle(
                    (i, ordered_data.shape[1]-1-j), 1, 1, linewidth=0, color=color_red
                )
            )
        elif ordered_data.iloc[i,j] == -1:
            ax.add_patch(
                plt.Rectangle(
                    (i, ordered_data.shape[1]-1-j), 1, 1, linewidth=0, color=color_grey
                )
            )

# offset y-axis ticks to center them
ax.set_yticks(np.arange(len(ordered_colnames))+0.5)
# need to reverse the order of the labels this time
ax.set_yticklabels(ordered_colnames[::-1], rotation=0)

# Create legend for main heatmap
main_legend_elements = [
    Patch(facecolor='white', edgecolor='black', label='Non-carrier'),
    Patch(facecolor=color_red, edgecolor='black', label='Carrier'),
    Patch(facecolor=color_grey, edgecolor='black', label='Missing'),
]
ax.legend(handles=main_legend_elements, title='Matrix', bbox_to_anchor=(1, 0.5), loc='upper left', frameon=True)

# Add secondary annotations
ax = make_axes_locatable(ax)

# Create a new axes for the secondary annotation
ax_fhr = ax.append_axes("top", size="7%", pad="2%")

# Define a colormap for the risk types
cmap_fhr = ListedColormap(['white',color_pink,color_red])

# Plot the 1D heatmap for risk types
values_fhr = df_fhr_ann_ordered['risk'].values

ax_fhr.imshow(values_fhr[np.newaxis,:], aspect='auto', cmap=cmap_fhr)

# Remove ticks
ax_fhr.set_xticks([])
ax_fhr.set_yticks([])

# Add y-axis label for secondary annotation
ax_fhr.set_ylabel('Risk', rotation=0, labelpad=5, va='center', ha='right')

# Create legend for FHR
legend_elements_fhr = [
    Patch(facecolor='white', edgecolor='black', label='SR'),
    Patch(facecolor=color_pink, edgecolor='black', label='GHR'),
    Patch(facecolor=color_red, edgecolor='black', label='FHR'),
]
ax_fhr.legend(handles=legend_elements_fhr, title='Risk', bbox_to_anchor=(1, -3), loc='upper left', frameon=True)

plt.title('Risk classification and constituent features')
plt.tight_layout()
fig.savefig('../fhr-heatmap-full-ann-by-risk.1May26.svg')

# %% [markdown]
Remove the constituent TP53 features

# %% [code]
# Remove intermediate features that are not directly used in FHR calling
cols_to_drop = ['TP53 Del','TP53 LoH','TP53 NS Mut','ISS III','Gain1q','Early relapse','Refractory']
df_fhr_abbrv_ann = df_fhr_ann.assign(**{
    'ISS III & Gain1q': lambda xs: xs.apply(lambda x: 1 if x['Gain1q']==1 and x['ISS III']==1 else 0 if (x['Gain1q']==0 or x['ISS III']==0) else -1, axis=1),
    'Early relapse/refractory': lambda xs: xs.apply(lambda x: 1 if x['Refractory']==1 or x['Early relapse']==1 else 0, axis=1)}).drop(columns=cols_to_drop)

# Create ordered data with covariates
covariates = ['risk']
df_fhr_ann_ordered = oncoplot_ordering(df_fhr_abbrv_ann, extra_covariates=covariates,prioritize_covariates=True)

ordered_data = df_fhr_ann_ordered.drop(columns=covariates)
ordered_colnames = ordered_data.columns

# Create OncoPrint with side axes
plt.clf(); 

fig,ax = plt.subplots()
fig.set_size_inches(12,4)

xlim = ax.set_xlim(0,ordered_data.shape[0])
ylim = ax.set_ylim(0,ordered_data.shape[1])

color_red = "#FF2222"
color_grey = "#CCCCCC"
color_pink = "#FFAAAA"

# instead of using axes.imshow, use axes.add_patch to draw rectangles
for i in range(ordered_data.shape[0]):
    for j in range(ordered_data.shape[1]):
        # draw a thin rectangle for each 1 in the data
        if ordered_data.iloc[i, j] == 1:
            ax.add_patch(
                plt.Rectangle(
                    (i, ordered_data.shape[1]-1-j), 1, 1, linewidth=0, color=color_red
                )
            )
        elif ordered_data.iloc[i,j] == -1:
            ax.add_patch(
                plt.Rectangle(
                    (i, ordered_data.shape[1]-1-j), 1, 1, linewidth=0, color=color_grey
                )
            )

# offset y-axis ticks to center them
ax.set_yticks(np.arange(len(ordered_colnames))+0.5)
# need to reverse the order of the labels this time
ax.set_yticklabels(ordered_colnames[::-1], rotation=0)

# Create legend for main heatmap
main_legend_elements = [
    Patch(facecolor='white', edgecolor='black', label='No'),
    Patch(facecolor=color_red, edgecolor='black', label='Yes'),
    Patch(facecolor=color_grey, edgecolor='black', label='Missing'),
]
ax.legend(handles=main_legend_elements, title='Matrix', bbox_to_anchor=(1.0, 0.6), loc='upper left', frameon=False)

# Add secondary annotations
ax = make_axes_locatable(ax)

# Create a new axes for the secondary annotation
ax_fhr = ax.append_axes("top", size="15%", pad="2%")

# Define a colormap for the risk types
# cmap_fhr = ListedColormap([color_grey,'white',color_pink,color_red])
# 1 May 26: remove grey as there is no longer the missing category
cmap_fhr = ListedColormap(['white',color_pink,color_red])

# Plot the 1D heatmap for risk types
values_fhr = df_fhr_ann_ordered['risk'].values

ax_fhr.imshow(values_fhr[np.newaxis,:], aspect='auto', cmap=cmap_fhr)

# Remove ticks
ax_fhr.set_xticks([])
ax_fhr.set_yticks([])

# Add y-axis label for secondary annotation
ax_fhr.set_ylabel('Risk category', rotation=0, labelpad=5, va='center', ha='right')

# Create legend for FHR
legend_elements_fhr = [
    Patch(facecolor='white', edgecolor='black', label='SR'),
    Patch(facecolor=color_pink, edgecolor='black', label='GHR'),
    Patch(facecolor=color_red, edgecolor='black', label='FHR'),
    # Patch(facecolor=color_grey, edgecolor='black', label='Missing'),
]
ax_fhr.legend(handles=legend_elements_fhr, title='Risk', bbox_to_anchor=(1.0, 0), loc='upper left', frameon=False)

plt.title('Risk classification and constituent features')

plt.tight_layout()

fig.savefig('../fhr-heatmap-abbrv-ann-1May26.svg')


# %% [code]
df_fhr_ann.to_csv('../fhr-annotations-raw.1May26.tsv',sep='\t')

# %% [code]
df_fhr_abbrv_ann.to_csv('../fhr-annotations.1May26.tsv',sep='\t')

# %% [code]
df_fhr_ann.iloc[:,:-1].apply(pd.Series.value_counts)

# %% [code]
df_fhr_abbrv_ann.apply(pd.Series.value_counts)
```
